# Large scale sequencing of SARS-CoV-2 genomes from one region allows detailed epidemiology and enables local outbreak management

**DOI:** 10.1101/2020.09.28.20201475

**Authors:** Andrew J. Page, Alison E. Mather, Thanh Le-Viet, Emma J. Meader, Nabil-Fareed Alikhan, Gemma L. Kay, Leonardo de Oliveira Martins, Alp Aydin, Dave J. Baker, Alexander J. Trotter, Steven Rudder, Ana P. Tedim, Anastasia Kolyva, Rachael Stanley, Maria Diaz, Will Potter, Claire Stuart, Lizzie Meadows, Andrew Bell, Ana Victoria Gutierrez, Nicholas M. Thomson, Evelien M. Adriaenssens, Tracey Swingler, Rachel A. J. Gilroy, Luke Griffith, Dheeraj K. Sethi, Dinesh Aggarwal, Colin S. Brown, Rose K. Davidson, Robert A. Kingsley, Luke Bedford, Lindsay J. Coupland, Ian G. Charles, Ngozi Elumogo, John Wain, Reenesh Prakash, Mark A. Webber, S. J. Louise Smith, Meera Chand, Samir Dervisevic, Justin O’Grady, The COVID-19 Genomics UK (COG-UK) consortium

## Abstract

The COVID-19 pandemic has spread rapidly throughout the world. In the UK, the initial peak was in April 2020; in the county of Norfolk (UK) and surrounding areas, which has a stable, low-density population, over 3,200 cases were reported between March and August 2020. As part of the activities of the national COVID-19 Genomics Consortium (COG-UK) we undertook whole genome sequencing of the SARS-CoV-2 genomes present in positive clinical samples from the Norfolk region. These samples were collected by four major hospitals, multiple minor hospitals, care facilities and community organisations within Norfolk and surrounding areas. We combined clinical metadata with the sequencing data from regional SARS-CoV-2 genomes to understand the origins, genetic variation, transmission and expansion (spread) of the virus within the region and provide context nationally. Data were fed back into the national effort for pandemic management, whilst simultaneously being used to assist local outbreak analyses. Overall, 1,565 positive samples (172 per 100,000 population) from 1,376 cases were evaluated; for 140 cases between two and six samples were available providing longitudinal data. This represented 42.6% of all positive samples identified by hospital testing in the region and encompassed those with clinical need, and health and care workers and their families. 1,035 cases had genome sequences of sufficient quality to provide phylogenetic lineages. These genomes belonged to 26 distinct global lineages, indicating that there were multiple separate introductions into the region. Furthermore, 100 genetically-distinct UK lineages were detected demonstrating local evolution, at a rate of ∼2 SNPs per month, and multiple co-occurring lineages as the pandemic progressed. Our analysis: identified a discrete sublineage associated with 6 care facilities; found no evidence of reinfection in longitudinal samples; ruled out a nosocomial outbreak; identified 16 lineages in key workers which were not in patients indicating infection control measures were effective; found the D614G spike protein mutation which is linked to increased transmissibility dominates the samples and rapidly confirmed relatedness of cases in an outbreak at a food processing facility. The large-scale genome sequencing of SARS-CoV-2-positive samples has provided valuable additional data for public health epidemiology in the Norfolk region, and will continue to help identify and untangle hidden transmission chains as the pandemic evolves.

**Major points:** In Norfolk and surrounding regions:

- 100 distinct UK lineages were identified.
- 16 UK lineages found in key workers were not observed in patients or in community care.
- 172 genomes from SARS-CoV-2 positive samples sequenced per 100,000 population representing 42.6% of all positive cases.
- SARS-CoV-2 genomes from 1035 cases sequenced to a high quality.
- Only 5 countries, out of 103, have sequenced more SARS-CoV-2 genomes than have been sequenced in Norfolk for this paper.
- Samples covered the entire first wave, March to August 2020.
- Stable evolutionary rate of 2 SNPs per month.
- D614G mutation is the dominant genotype and associated with increased transmission.
- No evidence of reinfection in 42 cases with longitudinal samples.
- WGS identified a sublineage associated with care facilities.
- WGS ruled out nosocomial outbreaks.
- Rapid WGS confirmed the relatedness of cases from an outbreak at a food processing facility.

## Introduction

In December 2019, a new coronavirus-related disease (COVID-19) was first reported in Wuhan, China (Huang et al. 2020); the causal agent was identified as the novel human coronavirus, SARS-CoV-2. Since then, SARS-CoV-2 has spread globally leading to 25 million confirmed infections and 850,000 deaths (as of 2020-09-02) (Dong, Du, and Gardner 2020). Two risk factors are associated with higher mortality: sex, as males are at higher risk than females; and age, as older age groups at substantially higher risk (Jin et al. 2020).

Whole genome sequencing provides high-resolution data that enables investigation of pathogen evolution and population structure (Bryant, Chewapreecha, and Bentley 2012). When combined with robust epidemiological data, it is possible to gain insights into SARS-COV-2 origins (Lu et al. 2020), transmission (both global [Alm et al. 2020] and local [Filipe et al. 2020]) and responses to control measures (Oude Munnink et al. 2020). Since the start of the pandemic, sequencing efforts and data sharing have facilitated tracking of the pandemic (Shu and McCauley 2017), identifying multiple independent virus introductions into different countries (Alm et al. 2020). The ability to assign identifiers rapidly to groups of samples that are related is essential in public health, as demonstrated for influenza (Smith et al. 2009). These identifiers can be formulated in different ways: from conserved sequences identified by multi-locus sequence typing (Page et al. 2017), by assigning SNP addresses (Dallman et al. 2018); or, in the case of SARS-CoV-2, through the assignment of lineages (Rambaut et al. 2020).

The COVID-19 Genomics UK (COG-UK) consortium (COG-UK 2020) is a UK-wide public health surveillance initiative comprising nearly 20 organisations from universities, research institutes and public health agencies, that was created to generate and analyse large-scale SARS-CoV-2 sequencing datasets to understand virus evolution, transmission and spread in the UK. These data allow a detailed insight into the course of the pandemic at the country, county and individual institution level. It was through large-scale analysis of SARS-CoV-2 genomes that evidence of a mutation (D614G) in the spike protein was revealed; it is likely that this mutation is responsible for increased transmissibility of the virus (Volz et al. 2020).

For the Norfolk region, we established a robust, rapid sequencing pipeline for SARS-CoV-2. Weekly sequencing data were fed back into the national effort for pandemic management, whilst simultaneously being used to assist local outbreak analyses. Here we describe the sequencing of genomes present in 1,565 SARS-CoV-2 samples from 1,376 cases, collected between March and August 2020. This represented 42.6% of all cases in the local population and included those with a clinical need, and key workers (such as healthcare, care and police) and their families. As of 2020-08-27, only 5 countries (UK, Australia, Spain, India and the USA) out of 103 countries have sequenced more SARS-CoV-2 genomes than have been sequenced in Norfolk for this paper. We used these data to investigate the genetic and epidemiological characteristics of the COVID-19 pandemic in the stable, low-density population of Norfolk and surrounding areas, UK. Our objectives were to use these sequence data to understand the evolution and spread of the virus locally, adding context to the national and global data, and to evaluate the role of rapid whole-genome sequencing for outbreak analysis in this setting.

Our analysis: identified a sublineage associated with 6 care facilities; found no evidence of reinfection in longitudinal samples; ruled out a nosocomial outbreak; identified 16 lineages in key workers which were not in patients indicating infection control measures were effective; found the D614G spike protein mutation which is linked to increased transmissibility dominates the samples and rapidly confirmed relatedness of cases in an outbreak at a food processing facility. This demonstrates the valuable role of large scale genome sequencing of SARS-CoV-2 to inform surveillance and regional outbreak management.

## Methods and Materials

### Samples

The clinical samples we used were initially collected passively for diagnostic testing with ethical approval from Public Health England (R&D ref NR0195) and with sampling directed by government public health policy and local clinical need. Samples were taken at four large hospitals: Norfolk and Norwich University Hospital (NNUH) (1,200 beds) in Norwich, Norfolk; The Queen Elizabeth Hospital (QEH) (500 beds) in Kings Lynn, Norfolk; The James Paget University Hospital (JPUH) (500 beds) in Great Yarmouth, Norfolk; and the Ipswich Hospital (550 beds) in Ipswich, Suffolk. Additional clinical samples that were included were collected at five smaller hospitals; by three community care organisations (representing dozens of care facilities and GP practices); and at drive through testing facilities for health care workers, essential workers (such as police) and their families who live or work in Norfolk and the surrounding areas.

### Samples and RNA extraction

Samples from cases with suspected SARS-CoV-2 were processed using five different diagnostic platforms over three laboratories on the Norwich Research Park - the Cytology Department and Microbiology Department, NNUH, Norwich, UK and the Bob Champion Research & Education Building (BCRE), University of East Anglia, Norwich, UK. Samples were primarily nasal/oropharyngeal swabs, although nasopharyngeal aspirates, bronchoalveolar lavage and sputum samples were also collected. The Cytology Department processed samples using the Roche Cobas® 8800 SARS-CoV-2 system (https://tinyurl.com/yy58t8sp). The Microbiology Department processed samples using either the Hologic Panther Fusion System SARS-CoV-2 assay (https://tinyurl.com/yye3m25p) according to the manufacturer’s instructions, the AusDiagnostics SARS-CoV-2, Influenza and RSV 8-well panel (https://tinyurl.com/yyeh5y2w) or Altona Diagnostics RealStar® SARS-CoV-s RT-PCR Kit 1.0 (https://tinyurl.com/y2avsrt9). RNA was extracted from swab samples in the Microbiology Department using either the QIAsymphony (Qiagen) or AusDiagnostics MT-Prep (AusDiagnostics) instruments according to the manufacturer’s instructions before being processed through the AusDiagnostics assay. In the BCRE, RNA was extracted using the MagMAX™ Viral/Pathogen II Nucleic Acid Isolation kit (Applied Biosystems) according to the manufacturer’s instructions and the KingFisher Flex system (ThermoFisher). The presence of SARS-CoV-2 was determined on either the QuantStudio 5 (Applied Biosystems) or Lightcycler LC480II (Roche) with the 2019-nCoV CDC assay (https://www.fda.gov/media/134922/download).

Viral transport medium from positive swabs (stored at 4°C) was collected for samples run on the Roche Cobas and Hologic Panther Fusion systems and in all other cases excess RNA was collected (stored at 4°C and collected within four days for samples tested by the AusDiagnostics assay, all other RNA samples were initially frozen and thawed for collection). Excess positive SARS-CoV-2 inactivated swab samples (200µl viral transport medium from nose and throat swabs inactivated in 200µl Zymo DNA/RNA shield and 800µl Zymo viral DNA/RNA buffer) were collected from Cytology and the Microbiology Department and SARS-CoV-2 positive RNA extracts (∼20µl) were collected from the Microbiology Department and the BCRE as part of the COG-UK Consortium project (PHE Research Ethics and Governance Group R&D ref no NR0195), with full details in Supplementary Tables 1-3. For inactivated swab samples, RNA was extracted using the Quick DNA/RNA Viral Magbead kit from step 2 of the DNA/RNA purification protocol (Zymo - https://tinyurl.com/y2lqoneq).

SARS-CoV-2 positive samples were transferred to the Quadram Institute Bioscience for sequencing. The lower cycle threshold (Ct) or take-off value produced by the SARS-CoV-2 assays in the Roche, AusDiagnostics, Altona Diagnostics and CDC assays were used to determine whether samples needed to be diluted for sequencing according to the ARTIC protocol (for AusDiagnostics results, 13 was added to the take-off value to generate an approximate Ct value - this is because 15 cycles of PCR are performed before a dilution step and a further 35 cycles of nested PCR (the take-off value is determined in the nested PCR)). The SARS-CoV-2 assay in the Hologic Panther does not provide a take-off or Ct value but rather a combined fluorescence signal for both targets in RLUs, therefore all samples tested by the Hologic Panther were processed undiluted in the ARTIC protocol.

### Sequencing using ARTIC SARS-CoV-2 multiplex tiling PCR

cDNA and multiplex PCR reactions were prepared following the ARTIC nCoV-2019 sequencing protocol v2 (Quick 2020). Dilutions of RNA were prepared when necessary based on Ct values following ARTIC protocol guidelines. V3 CoV-2 primers (https://github.com/artic-network/artic-ncov2019/tree/master/primer_schemes/nCoV-2019/V3) were used to perform the multiplex PCR for SARS-CoV-2 according to the ARTIC protocol (Quick 2020) with minor changes. Due to variable Ct values, all RNA samples used in the two ARTIC multiplex PCRs were run for 35 cycles. Odd and even PCR reactions were pooled and cleaned using a 1x SPRI bead clean with KAPA Pure Beads (Roche Catalogue No. 07983298001), according to the manufacturer’s instructions. PCR products were eluted in 30 µl of 10 mM Tris-HCL buffer, pH 7.5 and cDNA quantified using the QuantiFluor® ONE dsDNA System (Promega, WI, USA). Libraries were prepared for sequencing on the Illumina or Nanopore platform and sequenced as described previously (Baker et al. 2020).

### Sequence analysis

Raw reads were demultiplexed using bcl2fastq (v2.20) (Illumina Inc.) allowing for zero mismatches in the dual barcodes to produce FASTQ files. The reads were used to generate a consensus sequence for each sample using an open source pipeline adapted from https://github.com/connor-lab/ncov2019-artic-nf (https://github.com/quadram-institute-bioscience/ncov2019-artic-nf/tree/qib). Briefly, read adapters were trimmed using TrimGalore (https://github.com/FelixKrueger/TrimGalore) and aligned to the Wuhan Hu-1 reference genome (accession MN908947.3) using BWA-MEM (v0.7.17) (Li 2013); ARTIC amplicons were masked and a consensus built using iVAR (v.1.2) (Grubaugh et al. 2019).

### Quality Control

Samples were prepared and sequenced in 96-well plates with one cDNA negative control per plate and one RNA extraction negative control, where applicable. Contaminated samples were removed from analysis (Supplementary Table 2). The COG-UK consortium defines a consensus sequence as passing COG-UK quality control (QC) if: > 50% of the genome is covered by confident calls or there is at least one contiguous sequence of more than 10,000 bases; and no evidence of contamination in the negative control. This is regarded as the minimum amount of data needed to be phylogenetically useful. A confident call is defined as having 10x depth of coverage. If the coverage falls below these thresholds, the bases are masked with the character N indicating the base at that position is unknown or not available. Low quality variants are also masked with Ns. The QC threshold for inclusion in the public database GISAID (Global Initiative on Sharing All Influenza Data) is higher, requiring that > 90% of the genome is covered by confident calls and that there is no evidence of contamination. The COG-UK quality control criteria were used as the minimum requirements for lineage and phylogenetic analysis.

### Data availability

All consensus sequences were deposited in GISAID (Elbe and Buckland-Merrett 2017) if they met its minimum QC threshold. All raw sequence data and metadata (Griffiths et al. 2020) were deposited in the European Nucleotide Archive (ENA) (Cochrane et al. 2016). In both cases this happened soon after sequence generation, facilitated through COG-UK, and using MRC CLIMB (Connor et al. 2016).

### Clustering and phylogenetic analysis

Lineages (Rambaut et al. 2020) assigned to each consensus genome were determined using Pangolin (https://github.com/cov-lineages/pangolin) which is run routinely by the Rambaut group over SARS-CoV-2 consensus sequences deposited on MRC CLIMB (Connor et al. 2016). Global lineages are identifiers given to actively spreading lineages defined using a phylogenetic framework and often represent distinct introductions into new territories or regions, taking the form B.1.2.3, see (Rambaut et al. 2020) for full details. UK lineages represented the subsequent spread within the UK, taking the form UK1234, providing an identifier for a cluster for a given phylogeny, however the identifiers are not consistent between phylogenies. Only samples that passed COG-UK QC were considered for lineage assignment (>50% of genome reconstructed). Phylogenetic trees were visualised using ggtree (Yu 2020).

### Epidemiological analyses

Epidemiological analyses of outbreaks presented in the results were instigated and overseen by clinicians within the NHS or by public health bodies. Genome sequencing, bioinformatics analysis and genomic epidemiology was done by Quadram Institute Bioscience with limited anonymised metadata as part of the COG-UK consortium; patient-identifiable data were retained by the hospitals or public health bodies.

#### Longitudinal sampling and reinfection

For each case, the Norwich Research Park Biorepository (part of NNUH) anonymously linked all instances where cases were sequenced longitudinally and provided the information to Quadram Institute Bioscience for analysis. The UK lineages were extracted for each case with multiple samples using precomputed lineages from COG-UK. Consensus genomes which did not yield high enough quality genomes to compute a lineage were excluded. Cases which had more than 2 high quality samples were validated to ensure the lineages were the same. Where differences were identified, all consensus genomes for the case were extracted into a FASTA file and the differences compared to the Wuhan Hu-1 reference were noted using SNP-sites (Page et al. 2016) (version 2.3.3).

#### Care facilities outbreak analysis

An initial list of SARS-CoV-2 samples associated with a single care facility was provided by NNUH to Quadram Institute Bioscience. The UK lineages were identified for each sample using precomputed lineages from COG-UK. All other samples with the same UK lineage in the COG-UK dataset were identified and a phylogenetic tree computed using iQTree2 (Minh et al. 2020) (version 2.0.6). All samples in a sublineage associated with Norfolk were identified. The mutations defining this sublineage were calculated using SNP-sites (Page et al. 2016) (version 2.3.3), with the Wuhan Hu-1 reference as the base. A phylogenetic tree of the sublineage was calculated before first removing singleton mutations, most of whom where C->T/U SNPs, which are markers of RNA degradation. A phylogenetic tree was computed using iQTree2. Anonymised care facility sample metadata was added to the sublineage, with the data visualised in Phandango (Hadfield et al. 2018) and the relatedness of the samples and care facilities was visually confirmed. Hospital admission and discharge data for the residents was analysed solely by Public Health England co-authors and an anonymised summary was provided for this paper to maintain patient confidentiality.

#### Hospital outbreak analysis

A list of SARS-CoV-2 samples associated with a hospital were provided by Ipswich Hospital to Quadram Institute Bioscience. The lineages were identified for each sample using precomputed lineages from COG-UK. The frequency of each lineage was identified for the organisation. SARS-CoV-2 samples collected by other hospitals (NNUH, QEUH and JPUH) and care organisations had the frequencies of their lineages calculated in a similar fashion, providing context.

#### Food processing facility outbreak analysis

SARS-CoV-2 positive samples were sent to the Quadram Institute Bioscience. Samples were prepared, sequenced using an Oxford Nanopore Technologies MinION, and bioinformatically analysed, all within 24 hours of sample receipt. Consensus genomes were provided to Civet (https://github.com/COG-UK/civet), which assigned lineages to each genome. SNPs defining the sublineage of the outbreak were manually identified from the Civet report. The global lineage that the samples fell into was analysed further. All public samples from the global lineage which were publicly accessible through GISAID were identified. The countries of origin and collection dates were noted. A phylogenetic tree of all samples in this lineage from May onwards was created using iQTree2. The SNPs defining the sublineages were calculated using SNP-sites(Page et al. 2016) (version 2.3.3).

## Results

### Samples

The first reported case in the Norfolk region was on 2020-03-06 from a returning traveller; by 2020-08-31, there were 3,225 cases identified by NNUH from Norfolk and surrounding areas from a total of 3,751 SARS-CoV-2 positive clinical samples (some cases were sampled multiple times). Of these, 1,565 samples (41.7%) were sequenced and analysed, from 1,376 cases (42.6%). This represents approximately 172 SARS-CoV-2-positive samples sequenced per 100,000 population. The sequenced cases were broken down by locality and age group (Supplementary Table 4). For cases sampled multiple times, the earliest collection date of a SARS-CoV-2 positive sample was used for sequence analysis. These samples were collected in the East of England, predominantly from cases with providing an address in Norfolk (Supplementary Figure 1). The samples came from individuals in the community (20.7%, n=285), inpatients (40.6%, n=559) and outpatients (0.3%, n=4) at hospitals, and Staff (key workers) (23.8%, n=328) and their families (4.7%, n=65) (Figure 1). Inpatients represented a mixture of patients newly-admitted to the hospital (with or without COVID-19 symptoms), and existing patients with possible nosocomial SARS-CoV-2 infections. As testing was extended to more groups, so the regions from which samples were collected also changed (Figure 2).

**Figure 1:**
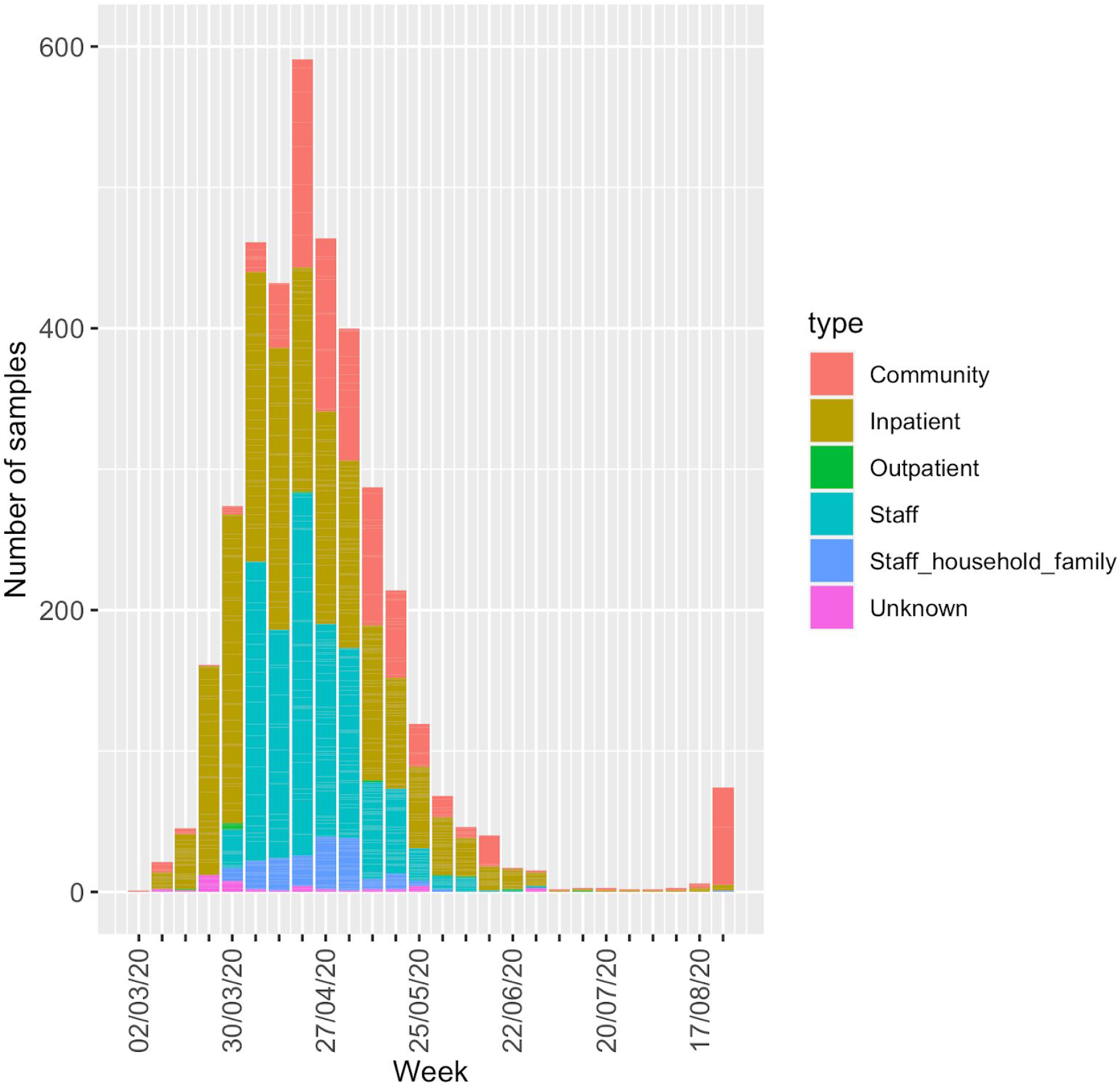
Total number of positive samples in the region per week, broken down by type. Not all of these were available for sequencing and a single individual may have been sampled multiple times. Staff (key workers) include healthcare workers and essential workers, such as police officers.

**Figure 2:**
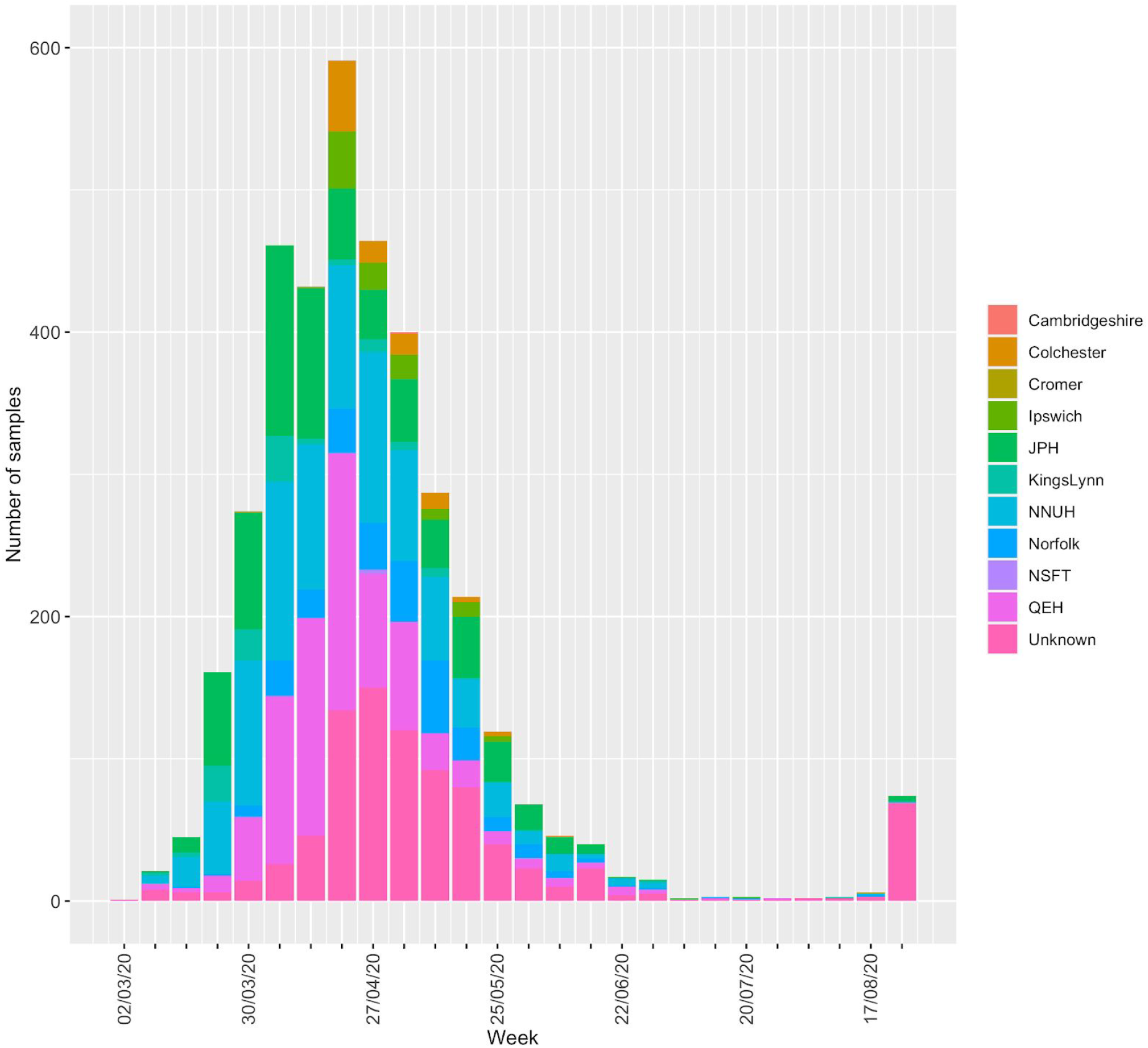
Total number of positive samples per week, broken down by the locality from which they were collected. If specific hospital information was not available, geographical location at the finest resolution was used. Not all samples were sequenced and a single individual may have been sampled multiple times.

The number of positive samples in the Norfolk region peaked at the end of April 2020; specifically, the number of SARS-CoV-2-positive samples peaked in the week of 2020-04-20 2020-04-26, with 591 positive samples (Figure 3). The peak month was April with 1,992 positive samples, followed by May with 1,188 positive samples. These numbers include a small number of repeat samples. Examining samples from new cases of infection only, in July 10 positive cases were reported, before rising to 79 in August. More than 60 of the August cases are related to food processing facility outbreak.

**Figure 3:**
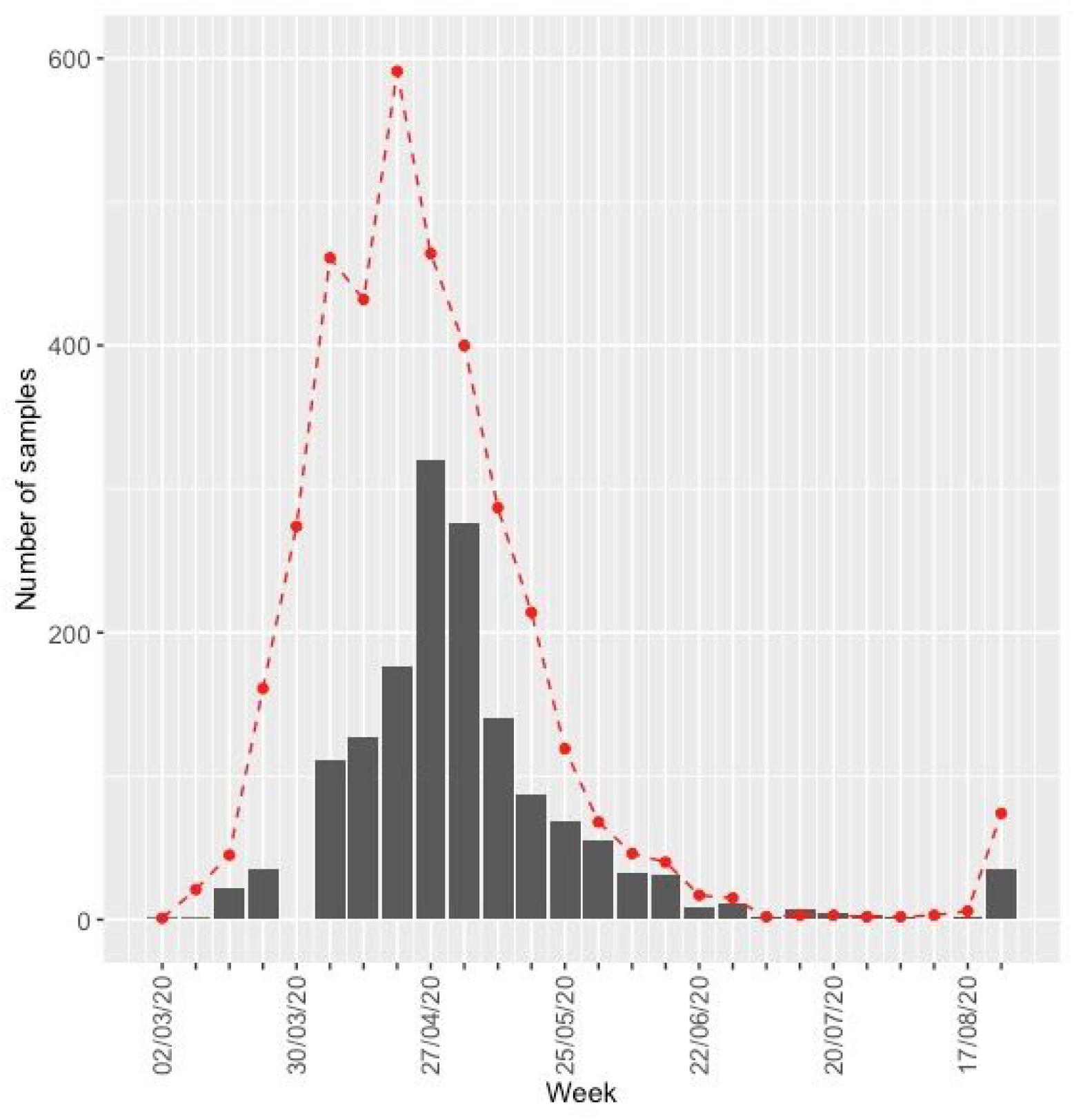
Number of positive samples sequenced at Quadram Institute Bioscience (grey bars) over time compared with the number of samples collected (red dashed line). Project sample collection only officially began on 2020-04-08 and not all samples taken before this time were available for sequencing. However, archived samples for March were sequenced and are represented in this figure.

Proportionally, the number of SARS-CoV-2-positive samples that were sequenced followed the same trend as the total number of positive samples, peaking in the week of 27 April to 3 May, with genomes from 320 cases being sequenced (Figure 3). Although project sample collection for sequencing officially began in 2020-04-08, and no samples were sequenced from the period 2020-03-27 to 2020-04-07, 59 archived samples from March were available and were sequenced. Overall the number of genomes sequenced does reflect the number of positive cases in the region and we can confidently conclude that the peak period was April/May 2020, in this region.

The number of positive cases sequenced was greatest in older individuals, with the largest number of samples (n=316, 36.5%) being from cases aged 80-90 years (Figure 4a). Just nine samples originated from cases under 10 years of age. Females were also significantly over-represented in the dataset (Figure 4b), accounting for 57% (n=741 out of 1286, p<0.001, one-proportion z-test) of cases. All samples made available for sequence were sequenced, with no selection criteria based on patient data or quality cut-offs, thus we expect the sequenced positive cases to proportionately reflect total positive cases.

**Figure 4:**
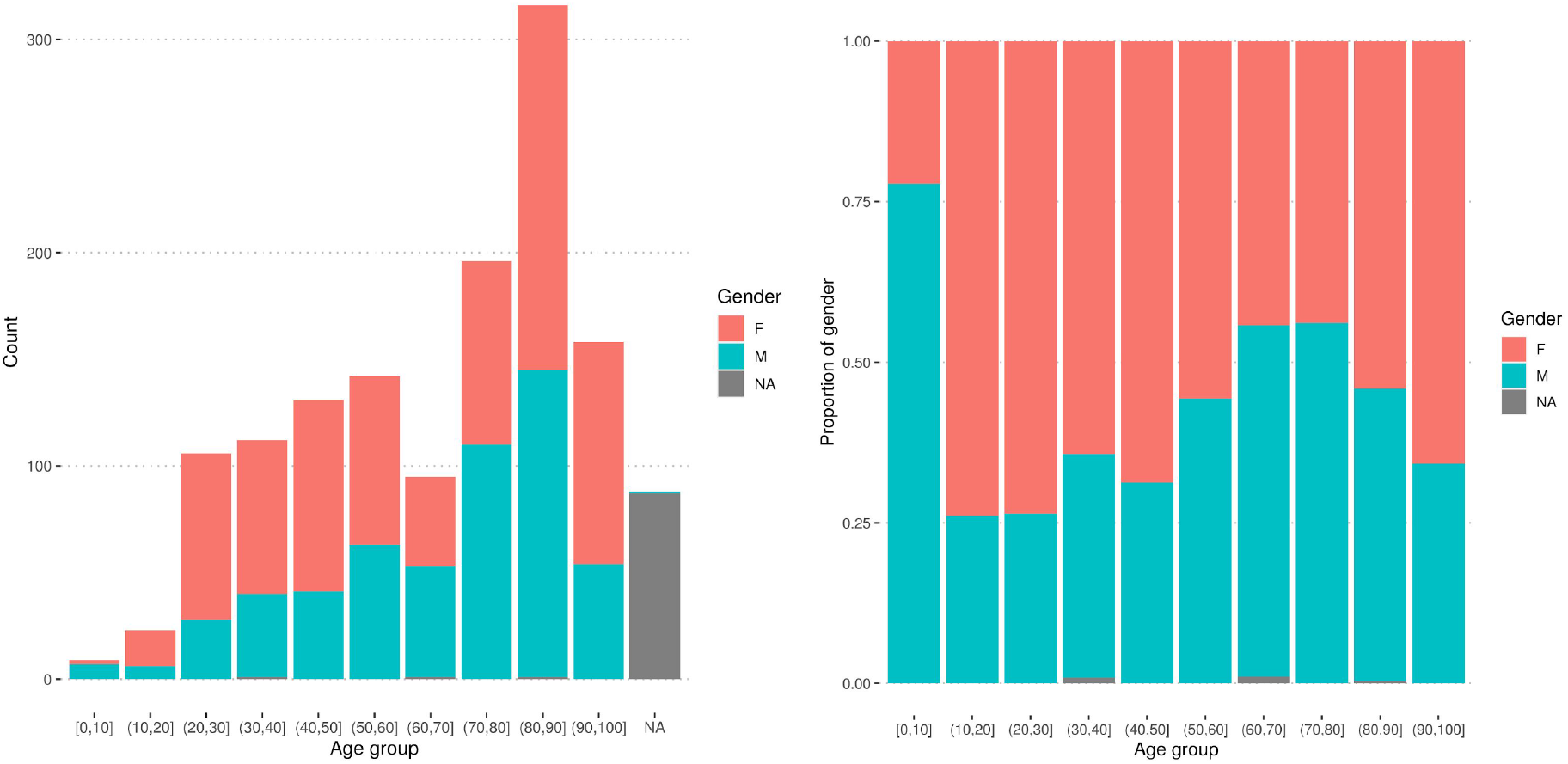
Age and sex of positive cases in absolute values (a) and scaled as proportions of all positive cases sequenced (b). Data from one person over 100 years of age is not included. NA: not available.

### Genomic Analysis

#### Genome quality control

Samples received for sequencing varied substantially in viral load. For our samples the Ct correlated well with the percentage of bases missing in the reconstructed consensus genome (Supplementary Figure 2) with a substantial reduction in genome completeness for samples with a Ct above 32. All diagnostic positives were sequenced, irrespective of Ct value, to avoid underrepresenting patients with low viral loads.

We identified potential biases in the quality of the genomes sequenced from samples. The sex and age of the cases (for those genomes which had the relevant associated metadata) were evaluated against four QC categories: not sequenced, failed all QC, passed basic QC, passed high quality QC (Supplementary Figure 3). There were significantly more genomes from females in each of the four QC categories (one-proportion z-test, p<0.001 for all but the not sequenced category [p=0.052]). With respect to an individual’s age, the mean age of individuals contributing samples that failed all QC was significantly higher (mean age 70.0 years vs 64.9, 65.7, 58.8 years of age of individuals contributing genomes passing basic QC, passing high quality QC, and not sequenced, respectively, p<0.05, pairwise Kruskal-Wallis rank sum tests adjusted for multiple hypothesis testing).

Completeness of consensus genomes is related to the Ct of the input samples (Supplementary Figure 4; Supplementary Figure 5) and three temperature-sensitive ARTIC PCR primer dropout areas are visible (Benjamin Farr et al. 2020) visible. A further three performed poorly at higher Ct values, due to reduced amplification efficiency, variation which is to be expected in a large amplicon pool. Overall, the ARTIC protocol was robust and sensitive. Above Ct 32 the completeness of the genomes recovered did begin to tail off and there was a substantial, largely random, drop off above Ct 35, i.e. there was no consistency in the primer pairs that performed well or poorly in the multiplex when there were < ten genome copies present.

#### Global lineages

As samples were collected over a six month period, the median number of SNPs per genome increased every month compared with the Wuhan Hu-1 reference (accession MN908947.3). When only considering high quality consensus genomes (Supplementary Figure 6) it increased from six SNPs in March to 16 SNPs in August. The evolutionary rate was estimated to be ∼2 SNPs per month.

The maximum number of co-occurring global lineages in a given week was 13 for the period 2020-04-27 to 2020-05-10, approximately 5-6 weeks after the UK government instituted a lockdown (2020-03-23) (Figure 5). This rapidly reduced as the number of samples dropped.

**Figure 5:**
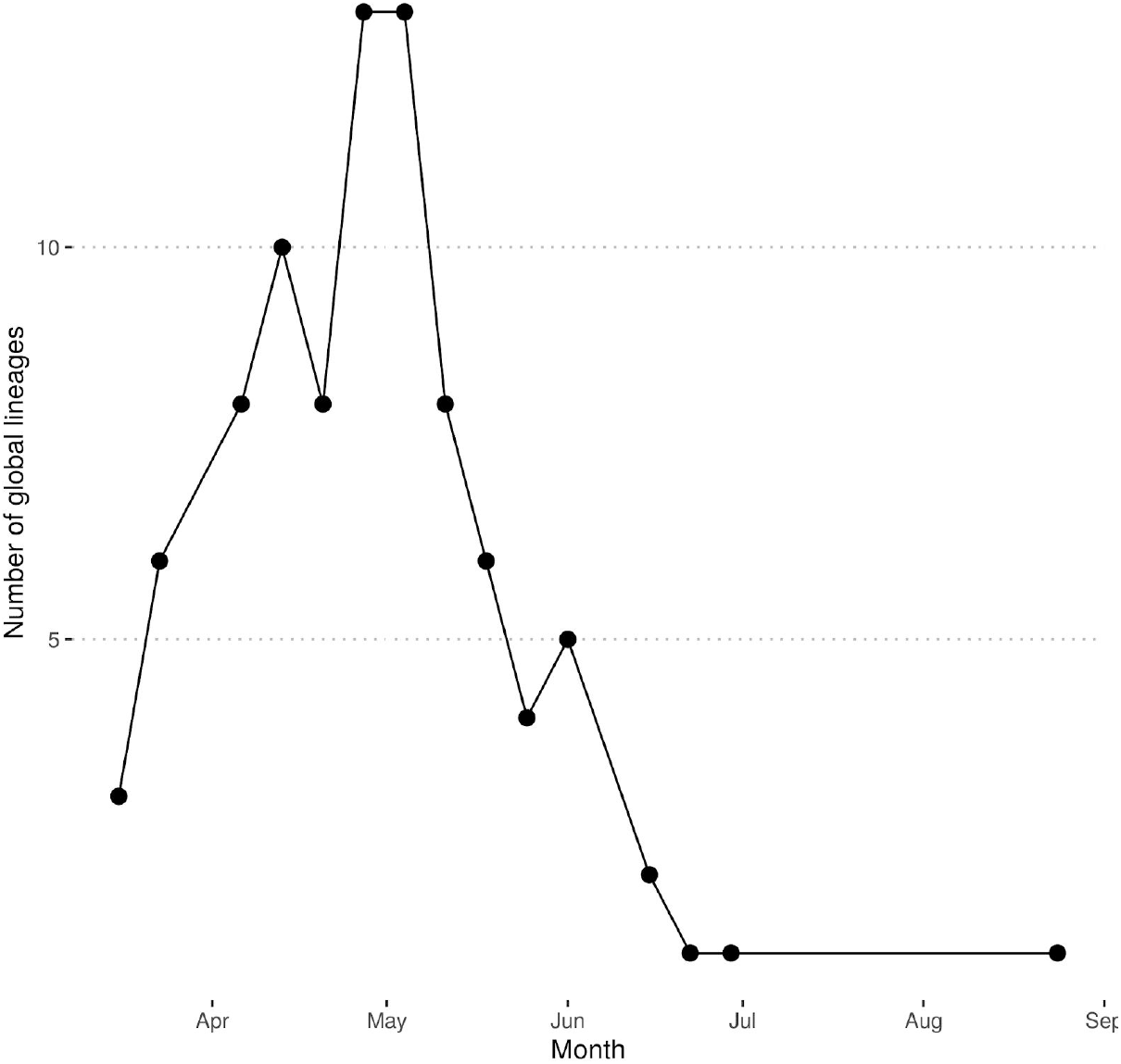
The weekly number of co-occurring global lineages, excluding lineages that were only found in a single sample.

When considering the number and proportion of co-occurring global lineages every week it is apparent that, during the peak (April/May), some global lineages became extinct and were replaced by new lineages, which rapidly increased in abundance (Figure 6). The transient nature of lineages is exemplified in lineages B.1.1.3 and B.2. Lineage B.1.1.3 was first identified in the week beginning 2020-05-04 (seven cases); it persisted for a month before becoming extinct within the region. Lineage B.2 was observed much earlier, in the week beginning 2020-03-16 (two cases), peaked one month later (19 cases) and was last observed on 2020-05-11 (two cases).

**Figure 6:**
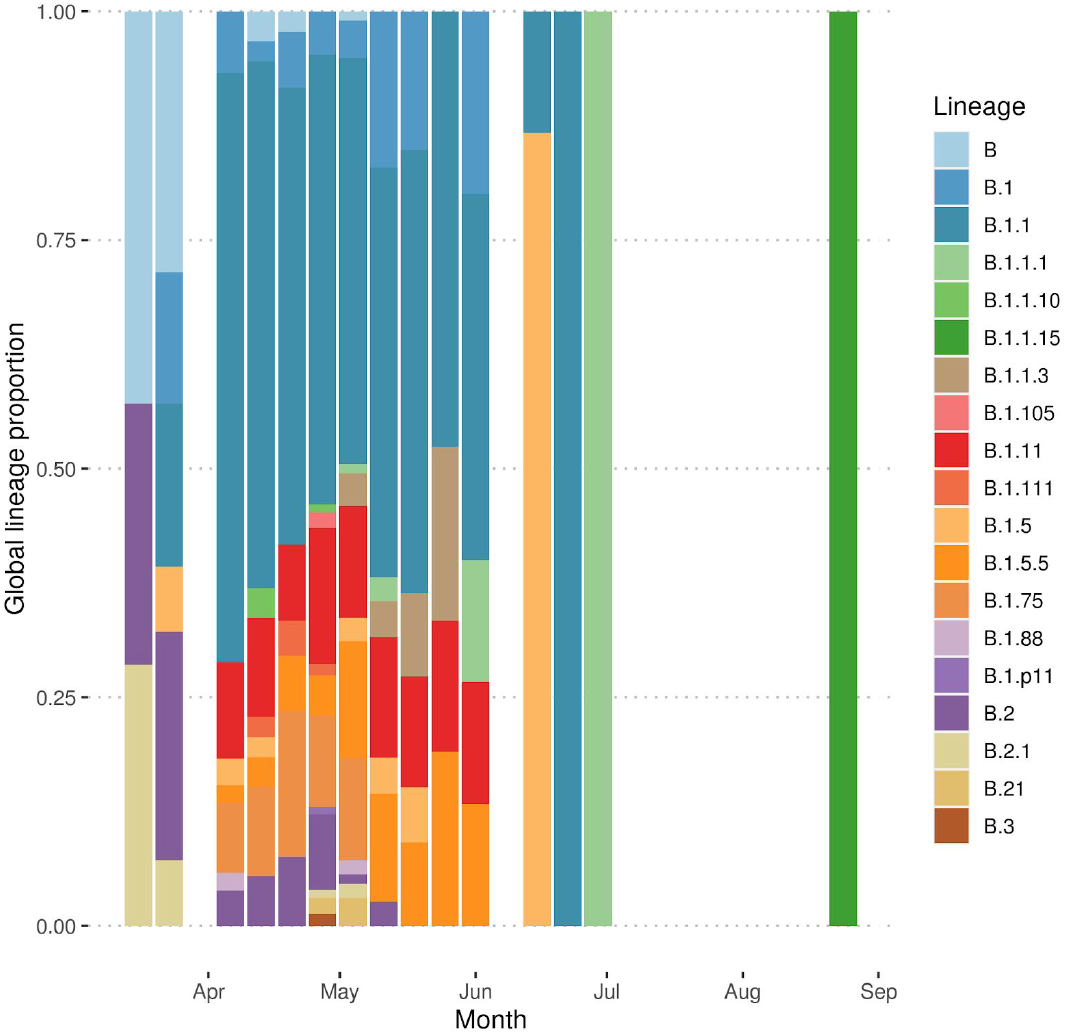
The proportion of samples represented by each lineage per weekly, excluding lineages represented by a single sample. Sample collection for sequencing began on 2020-04-08, and no archival samples were available for sequencing for the period 2020-03-26 to 2020-04-07. Samples from July and August were primarily repeated sampling of the same case or with only a single individual in a lineage.

The global lineage nomenclature system of Rambaut et al. (2020) provides a flexible and consistent naming scheme for genomically-detectable introductions of SARS-CoV-2 into new locations; there have been 1035 global lineages assigned in this scheme. A total of 26 of these global lineages were observed in our data, 20 of which were observed in more than one sample. All of the global lineages present in more than one sample were from lineage B, which is predominantly associated with European and UK outbreaks (https://github.com/hCoV-2019/lineages/blob/master/lineages/data/lineage_descriptions_2020-05-19.txt). A number of lineages were over- or under-represented in Norfolk samples compared with the UK and globally (Table 1). For example: B.1.1.15 was represented in 3.38% (n=35) of Norfolk genomes compared with 0.97% (n=60) globally (only 25 from outside Norfolk); B.1.11 which was represented in 10.72% of Norfolk genomes compared with 1.34% globally; B.1.5.5 which was represented in 6.47% of Norfolk genomes compared with 0.12% globally; and B.1.75 which was represented in 8.12% of Norfolk genomes compared with 0.25% globally.

**Table 1:**
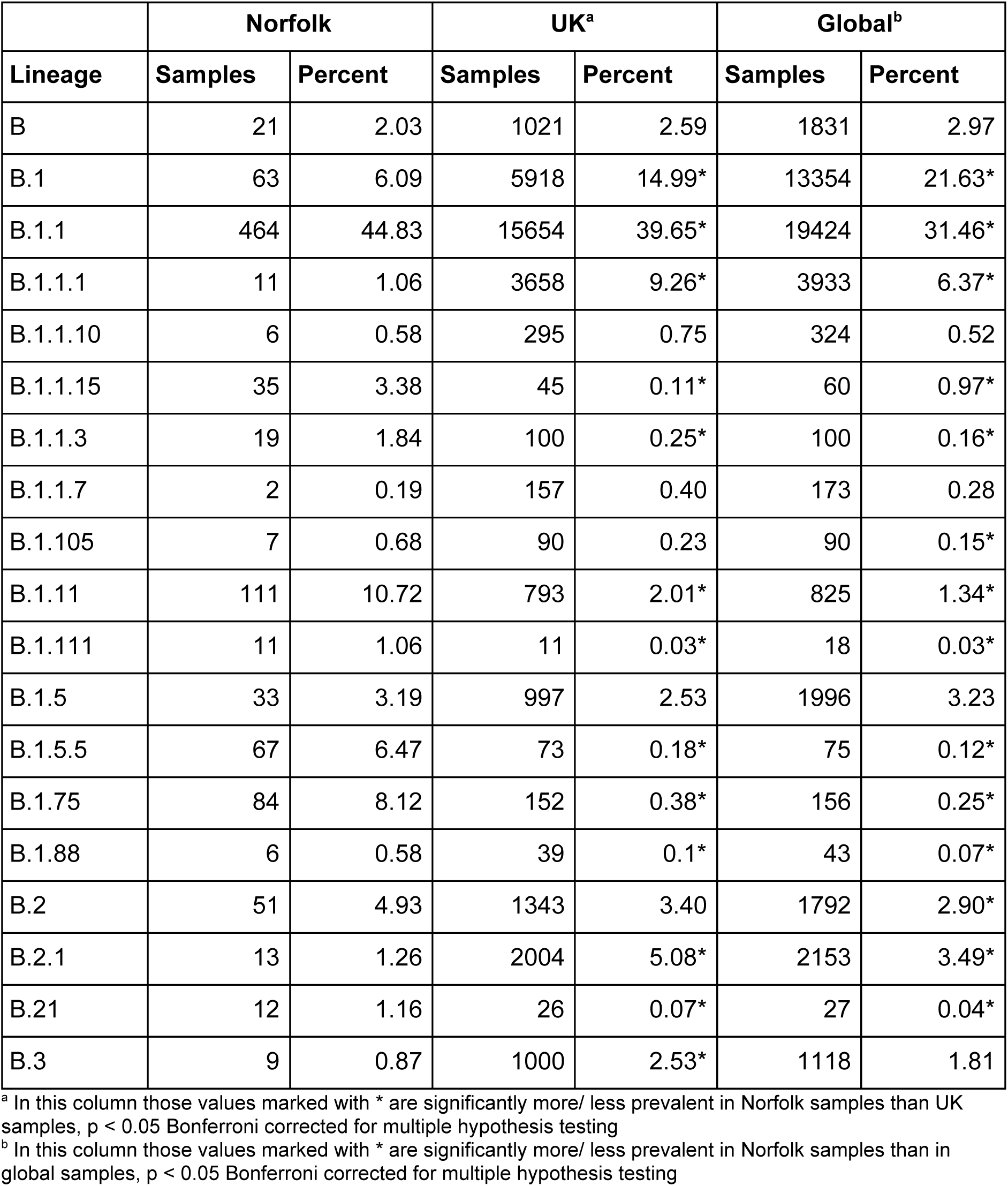
Summary of the prevalence of global lineages represented by more than one sample in the Norfolk dataset compared with the public set of genomes from GISAID and COG-UK (accessed 2020-08-28).

The most commonly sequenced global lineage was B.1.1 which was present in 464 samples (44.8%) in this dataset and 19424 globally (31.4%); this lineage has three defining single nucleotide polymorphisms (SNPs) at 28881 G->A, 28882 G->A and 28883 G->C and is closely associated with the European expansion of SARS-CoV-2.

Norfolk samples were set in context as part of the COG-UK phylogenetic pipeline (2020-09-07) using a phylogenetic tree based on all publicly-released genome sequences (Figure 7). Overall genomes from the Norfolk region represented a random sampling of co-occurring global lineages within the UK as a whole. Some major global lineages are under-represented in Norfolk such as B.1 and others are over-represented, such as B.1.1, when compared with the UK samples (Table 1).

**Figure 7:**
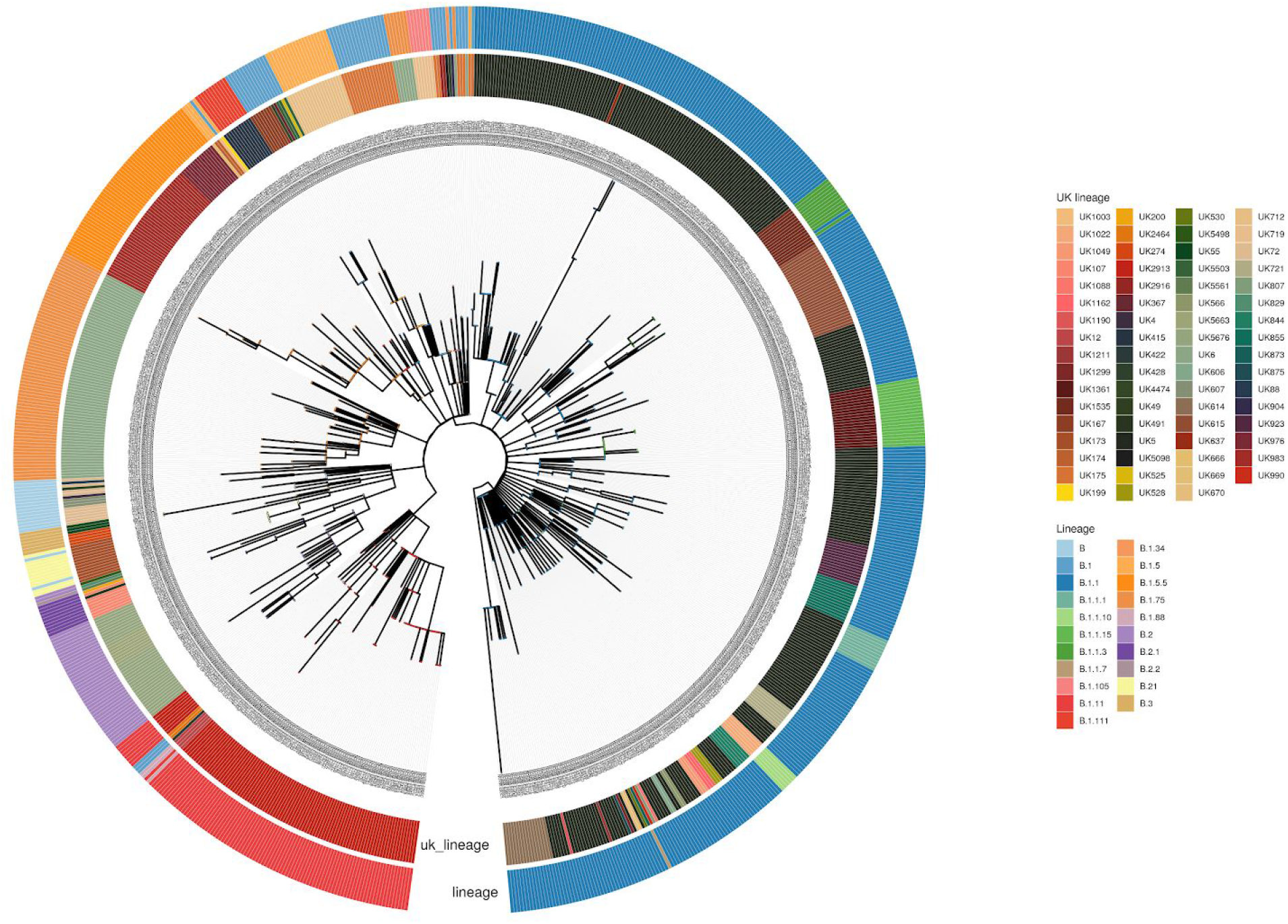
Phylogenetic tree of the Norfolk genomes sequenced in this study. The phylogenetic tree was estimated as part of the COG-UK phylogenetic pipeline (2020-09-07). The inner circle represents the UK lineages assigned to each sequence, while the outer circle shows their equivalent global lineages. Only high-quality samples are included (838 sequences).

#### UK lineages

Global lineages were further subdivided into UK lineages, to identify ongoing transmission and evolution within the UK. The numbers assigned to UK lineages are subject to change and must be recalculated for all genomes with each phylogenetic reanalysis. Thus the numbers reported here are for a single phylogenetic analysis.

There were 100 UK lineages detected in the dataset, 49 of which were present in two or more cases. The number of co-occurring UK lineages peaked at 20 in the week of 2020-04-27, approximately five weeks after the UK nationwide lockdown began; thereafter the number dropped to a single lineage in July and August (Supplementary Figure 7). The proportions of samples with particular lineages varied week to week (Supplementary Figure 8), with the most common UK lineage being UK5 which was present in 324 cases; this is also the most commonly identified lineage in the UK (https://microreact.org/project/cogconsortium-2020-09-02/f5aa0bdd/). The next most common UK lineage was UK2913 which was present in 113 samples; this was a sublineage associated with care facilities in the region around Norwich city (detailed later).

#### Examination of the D614G mutation in the spike protein

There is evidence that a mutation in the spike protein of SARS-CoV-2 (an amino acid change from D to G at position 614; D614G) increases infectivity of a pseudotype virus in vitro in cells; this is associated with an observed increase in viral loads in patients (Korber et al. 2020). Overall, in the Norfolk dataset, 89.4% (n=819) of samples had the D614G mutation while only 10.6% (n=97) had the wild type (Figure 8). The relative proportion of the two genotypes differed over time. In March, 66.6% (n=24) of samples contained the wild type and 33.3% (n=12) contained the D614G mutation. In April the proportion of genomes that were wild type had reduced to 10.7% (n=47) while those with the D614G mutation were dominant at 89.3% (n=392). In May the proportion of genomes that were wild type had reduced to 5.5% (n=22) compared with 94.4% (n=374) of genomes having the D614G mutation (Figure 8).

**Figure 8:**
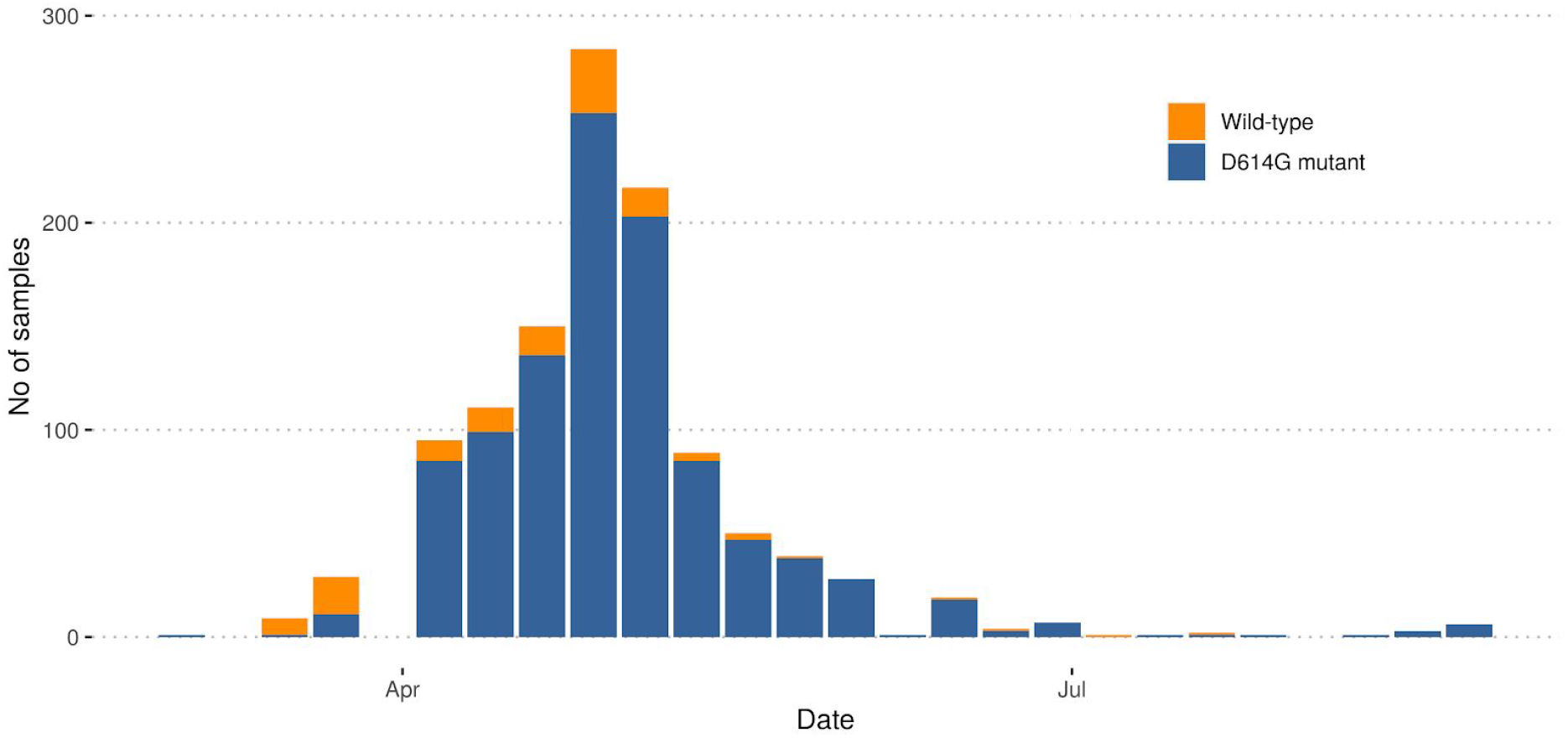
Weekly numbers of genomes with the wild type genome (D614) or the D614G mutant in the spike protein.

### Epidemiological Case Studies

#### Longitudinal sampling and reinfection

In August 2020 Quadram Institute Bioscience, at the request of the Microbiology Department at the NNUH, evaluated the genomes present in multiple longitudinal samples taken from the same case over extended periods of time during infection. The aim was to determine whether they were infected by the same lineage or different lineages, the latter indicating potential reinfection. Longitudinal SARS-CoV-2 positive samples were identified from 140 cases (Supplementary Table 3), each with between 2-6 samples, 88.5% of whom were hospital inpatients at some point during their illness. The median time span of the sampling was 13 days, with a mean of 16.2 days. The longest time span was 71 days, with 22 cases having a time span greater than 28 days. The clinical outcomes were not available for analysis. Only samples with different collection dates from the same individual were considered. We limited cases to those with high quality consensus genomes (passing GISAID QC) in two or more of the sample timepoints; this resulted in a series of longitudinal genome samples from 42 cases; each series had 2-4 samples. In every example the lineage remained the same between samples from the same individual, with the exception of the linked samples NORW-ED449 and NORW-ECD30; this was because NORW-ECD30 had nine IUPAC (IUPAC-IUB Comm. on Biochem. Nomenclature (CBN) 1970) symbols for ‘partially’ ambiguous bases, which are likely to be due to differences in viral load between the original samples (Ct 17 vs Ct 27). These results suggest that there is no evidence of reinfection in any of the individuals for which a series of positive samples had been taken.

#### Hospital outbreak analysis

In August 2020 Quadram Institute Bioscience and Ipswich Hospital used the genome data from a set of 31 samples from hospital patients to determine whether these samples represented a single nosocomial outbreak or whether they were unrelated and the result of community transmission. The 31 positive samples were collected between 2020-03-06 and 2020-07-28; 80.6% (n=25) were from patients over the age of 65. From these, 18 yielded genome sequences of sufficient quality to assign lineages. A total of six global lineages and eight UK lineages were observed in the samples, with the most commonly observed (n=5) being UK5, which is also the most commonly observed lineage within the UK. This number of co-occurring lineages indicated that there was not a single large nosocomial outbreak at this location.

#### Care facilities outbreak analysis

In June 2020 the Microbiology Department at NNUH and Quadram Institute Bioscience evaluated an outbreak at a care facility in the Norwich region using SARS-CoV-2 sequenced genomes from the dataset in this paper. The analysis undertaken indicated likely intra-care facility transmission, corresponding to a discrete sublineage circulating in the care sector as opposed to the wider community. It revealed that 14 out of 15 genomes from cases had the same UK lineage, UK2913, over a sustained period of time (2020-04-08 - 2020-06-01). An analysis of this lineage in all COG-UK data (n=395) revealed that it represented a distinct sublineage in the Norwich region of Norfolk, defined by a single synonymous mutation (A->G) at position 24232 in the S gene; this mutation was not found in any other COG-UK lineages. Most of the cases with this sublineage were >80 years of age and concentrated in distinct areas in the Norwich region. Analysis also confirmed that the samples were predominantly collected from six care facilities. There were 89 cases sequenced in the Norwich region with this sublineage and 76 of these were known to be patients (n=64) of care facilities or healthcare workers in those facilities (n=9) and their families (n=3). Links could not be established for 13 cases who tested positive for this sublineage. This sublineage had not been observed previously in community testing and the last new positive patient with this sublineage was on 2020-06-01. As it has not been seen in three months this sublineage is now regarded as extinct.

An analysis was undertaken to understand the role of hospital discharges in this sublineage. Of the residents in this sublineage, 12 had a hospital admission, of whom two were admitted twice to three hospital trusts. Six had a community-acquired infection, testing positive within seven days of admission, three were inconclusive due to missing data, one had a probable hospital-acquired infection and tested positive within seven days of discharge, and two had a definite hospital-acquired infection (https://www.gov.uk/government/publications/wuhan-novel-coronavirus-infection-prevention-and-control/epidemiological-definitions-of-outbreaks-and-clusters-in-particular-settings). All residents with a hospital-acquired infection had a test prior to discharge, suggesting the package of IPC measures were being followed. In the time period covered by the study patients required a test prior to discharge to a care facility, and a positive test did not preclude them from returning to the care facility, bur rather than enhanced IPC and isolation measures needed to be taken for a designated period of time. Given that some of this cohort of patients tested positive in May with community-acquired infections, a number of weeks after the adult social care IPC Department of Health and Social Care measures were announced, suggests that these measures may not have been sufficient.

On examination of all genome sequences obtained from a town with two of these care facilities, we found 70 samples were positive for SARS-CoV-2, with 52 of those yielding genome sequences of sufficient quality to assign a lineage. Thirty seven samples (71%) were associated with the care facilities and were the UK2913 lineage. The remaining 15 samples in the town came from 13 different lineages, indicating that the number of co-occurring lineages within the care facilities didn’t reflect the number of co-occurring lineages within the wider locality.

#### Food processing facility outbreak analysis

In August 2020 35 positive samples from workers at a food processing facility were rapidly sequenced (< 24 hours) as part of an outbreak analysis by Quadram Institute Bioscience, the Specialist Virology centre at NNUH, and Norfolk County Council Test and Trace and Public Health England. A total of 27 yielded genomes of sufficient quality to assign lineages. All genomes fell within the same cluster with a global lineage of B.1.1.15 and a UK lineage of UK1361, confirming that these genomes were related to each other. The consensus sequences of the genomes in this outbreak had three SNPs defining a sublineage: U/T->C at position 7783, G->U/T at position 25552 and G->U/T at position 28221. This sublineage had not been observed before in the Norfolk data, COG-UK data or global GISAID data.

Looking more widely at the lineage, it was only observed in samples from the UK, with seven other samples identified by community sequencing in July (2020-07-15 to 2020-07-27). However these seven samples lacked the three sublineage defining SNPs found in the outbreak samples, and had two additional defining SNPs of their own (G->U/T at position 12067, G->U/T at position 28086). This indicates that these two sets of samples had a common ancestor and, based on an evolutionary rate of ∼2 SNPs per month, this ancestor must have occurred 1-2 months earlier. Ancestors of lineage B.1.1.15 were only observed in Portugal in May and June (GISAID, accessed 2020-08-27).

## Discussion

Here, we have used intensive whole genome sequencing of SARS-CoV-2 samples in a single geographical area to investigate the evolution and transmission of the virus in this region. The average age of the Norfolk population is significantly higher than that of England as a whole; 24.5% of residents are aged 65 or older compared with 18.4% for England as a whole (https://www.norfolkinsight.org.uk/population/, accessed 31/05/20). The largest hospital in the region is the Norfolk and Norwich University Hospitals NHS Foundation Trust (1,200 beds), serving a population of around one million patients from Norfolk and neighbouring counties, supported by a network of smaller hospitals.

There was a lower incidence of SARS-CoV-2 in the Norfolk region compared with England as a whole, with the proportion of cases testing positive for SARS-CoV-2 in Norfolk at 363.2 per 100,000 compared with 573.9 per 100,000 in England as a whole (accessed 2020-09-14 https://coronavirus.data.gov.uk/). This was also reflected in the number of deaths that occurred due to infection within 28 days of a positive diagnosis; for Norfolk this was 43.8 per 100,000 population compared with 65.7 per 100,000 for England as a whole, which is significantly below average despite having an older, more vulnerable population.

The sequencing data represented a rate of 172 sequenced genomes per 100,000 in the Norfolk population, which corresponded to 113.8 cases for which high quality genome sequences were available for evaluation per 100,000 in the population. Specifically we evaluated high quality genome sequence for 31.3% of all cases that tested positive for SARS-CoV-2 in Norfolk. However, as the samples primarily came from health care settings, they captured cases with the most severe symptoms rather than asymptomatic community cases. Only positive tests from the population with clinical need (primarily hospital and care facilities), and key workers and their families were available for sequencing. Community testing was done at large regional ‘Lighthouse labs’ capable of processing tens of thousands of samples per day, with genome sequencing managed by the Wellcome Sanger Institute. Of the sequenced community samples just 25 attributed to Norfolk and lacked metadata, thus were not included in analysis.

The highest number of positive samples in this region occurred at the end of April/ beginning of May 2020, approximately six weeks after the UK instituted a nationwide lockdown (2020-03-23). Thereafter, the number of positive cases we sequenced dropped substantially as the impact of the lockdown and social distancing began to reduce transmission. With the exception of the food processing facility outbreak, by August 2020, only two new cases with positive samples were detected.

Analysis of the demographic metadata associated with positive samples indicated cases were more likely to be older and female. This skewed distribution in relation to age and sex is likely to be due to the directed use of diagnostic testing to symptomatic cases during the peak of the pandemic; this approach was driven by global shortages in reagents and testing capacity. Thus vulnerable elderly cases were more likely to be tested during the peak of the pandemic, and were more likely to be female as they have a longer life expectancy. Thereafter when testing was opened up to key workers, they were predominantly female healthcare workers who make up 77% of the NHS workforce (NHS 2019).

Viral loads in individuals, as measured by PCR, were correlated strongly with the percentage of the genome that could be reconstructed from the sequencing data. This could be due to individual variation in host factors, disease stage (Zou et al. 2020) or quality of the sample material (Rogers et al. 2020). Phylogenetically useful genomes where the Ct was below 32 (more than ∼100 viral copies) were routinely sequenced but there was a substantial tail off from Ct 35. These results must be interpreted cautiously in terms of transmission potential, as it is not known whether an individual is infectious at low viral loads; it is possible that the positive results from high Ct samples are due to detection of residual RNA from a past infection. However, this does demonstrate that sequencing can produce usable information from samples containing the wide range of viral loads likely to be encountered during SARS-CoV-2 infections.

There is evidence that a mutation in the spike protein with an amino acid change of D to G at position 614 (D614G) increases the transmissibility of the virus, which is associated with an increased viral load in mutant-infected cases (Korber et al. 2020; Volz et al. 2020). This has been observed in the UK and globally (Volz et al. 2020), potentially indicating that a more transmissible strain is now in circulation. As seen in Figure 8, in March 2020, during the early part of the pandemic in the Norfolk region, the wild type was found in the majority of samples (66.6%), although the overall number of sequenced samples were low. However there was a rapid fall in the proportion of samples with the wild type in April (10.7%), and by May 95% of samples had the D614G mutation. This corresponds to a changing landscape of global lineages in the region, with an increasing dominance of B.1 lineages and sublineages, and a decreasing occurrence of other B (non B.1) lineages.

The information provided by these sequences allowed an examination of the overall genetic variation within SARS-CoV-2 circulating in Norfolk and comparison with other regions. The number of co-occurring global lineages was similar to the range found within the UK as a whole (Filipe et al. 2020), Europe (Alm et al. 2020) and beyond (van Dorp et al. 2020). The notable exception was the lack of lineage A samples within the region, with only two being observed. This indicates that most of the lineages that entered the region did not come directly from China; rather, they are estimated to have predominantly come from Europe or within the UK. In the region, 23.2% (n=26) of all global lineages were observed out of 112 lineages that have been defined to date (Rambaut et al. 2020). This variation shows that genomically distinct lineages have expanded world wide, with different distributions taking hold in different settings (see Microreact [Argimón et al. 2016] https://microreact.org/project/cogconsortium-2020-09-02/f5aa0bdd/). The B.1.11 samples in our dataset were specifically associated with care facilities in Norfolk.

These data demonstrate a substantial number of co-occurring global lineages within one small region, indicating multiple concurrent introductions and their subsequent spread. This places a lower bound on the number of independent introductions to the region at 27, but it is likely to be substantially higher as not all COVID-19 infections were identified, tested and sequenced. As case numbers rose during the course of the pandemic, more lineages were identified, with a peak in the number of co-occurring lineages around five to six weeks after the UK instituted a national lockdown. Thereafter the number of lineages dropped substantially, with many rapidly becoming extinct in the region, providing further evidence that lockdown measures break transmission.

By subdividing the global lineages into standardised UK lineages, a finer resolution of viral genomic relatedness was obtained and allows us to make more detailed comparisons. As we observed with global lineages, the UK lineages provided further evidence for substantial viral genomic variability circulating in the region. There were 100 UK lineages observed in the region out of a total of 1725 lineages reported for the UK as a whole (5.7%). The dominant lineage in the UK (UK5) was also the dominant lineage in the region. Interestingly, the second most commonly observed lineage in the UK (UK1535), with 2269 (5.7%) samples and widespread circulation around the UK, was only observed 19 (1.8%) times in Norfolk (18 in this dataset, one sequenced by the University of Cambridge and none in Pillar 2 community testing for Norfolk).

One of the most important applications of these data was in epidemiological investigations to identify outbreaks. This was particularly important during the peak in April and May, when the number of new infections was high, providing the resolution required to distinguish between transmission clusters that would not have been otherwise possible. Our genomic data were informative in the following cases:

1. In a hospital setting, lineage information was used to differentiate nosocomial from community transmission. We sequenced 31 samples from Ipswich Hospital and founf eight UK lineages; the most commonly observed (UK5) was also the most commonly observed in the UK. This demonstrated that a single large nosocomial outbreak had not occurred but that the patients in hospital had become infected in the community by circulating lineages.
2. Our data unexpectedly uncovered a sustained outbreak in six care facilities within the region. This data indicated likely intra-care facility transmission and is currently under further investigation. The outbreak was identified while looking at a common lineage within the region and noticing that the positive samples had mostly come from elderly people, suggesting a possible link to care facilities. Further investigation at NNUH, identified six care facilities sharing a distinct sublineage primarily found only in these facilities. This sub-lineage was not detected in community testing (Pillar 2) at any point. Only 2 cases were a definite hospital acquired infection with 1 probable hospital acquired infections. Examination of all genome sequences obtained from a town with two of these care facilities, showed there were 13 different lineages circulating within the locality, but only a single lineage circulating in the care facilities.
3. Rapid genome sequencing was applied to an outbreak in a food processing facility in the region, yielding lineages for 27 genomes in 24 hours. Sequencing this subset of samples (about 25% of positive cases detected) identified virtually identical genomes that were not found in the general community prior to the outbreak. This demonstrated within factory transmission of a new lineage. An additional G->A mutation at position 20125 found in three of the outbreak genomes, demonstrated that within the short period of time that the virus was circulating in the factory, some evolution with subsequent onward spread had occurred. As the lineage had not been seen in the region previously, we are currently monitoring for transmission from the factory into the community. The factory sub-lineage is sufficiently novel to link it to the factory if identified anywhere in the country. Looking more widely at all publicly available data in GISAID, the only other country in which this global lineage (B.1.1.15) was recently observed was Portugal between 2020-05-06 and 2020-06-14 (the most recently observed genome from Portugal was from 2020-06-22). Genomics can be used to identify the possible source of a new introduction into a new area.

Samples could be broken down by the areas from which the samples were collected, representing towns or cities and their surrounds (Figure 2; Supplementary Figure 9). Whilst UK5 was the commonest UK lineage in most urban areas in the Norfolk region, as it was nationally in the UK, there were other UK lineages that showed sustained persistence and spread within discrete communities from small geographical areas. UK2913 was primarily observed in Norwich and to the south and east of the city (South Norfolk, Waveney, Broadland, Great Yarmouth), nearly exclusively from clients of care facilities and health care workers at those facilities. Another UK lineage, UK721, was observed in seven community care residents (aged 78-92) and two healthcare workers in south west Norfolk. One lineage, UK173 was observed only in one suburb of Norwich it dominated for one month (2020-04-13 to 2020-05-19), then went extinct and has not been observed in Norfolk since. This indicates that lineages introduced into small urban areas expand but are quickly halted by effective public health measures. The UK6 lineage was primarily observed in the Kings Lynn area, accounting for 90% (73/81) of UK6 samples in Norfolk. In contrast Norwich, 70 km away, recorded only a single case of UK6. These patterns are repeated throughout the dataset.

The number of co-occurring global and UK lineages in circulation amongst inpatients (n=559, 22 global, 71 UK) was also reflected in key workers and their families (n=394, 19 global, 49 UK). Sixteen UK lineages were observed in key workers and their families but were not observed in patients or in community care, though it must be noted that 10 of these lineages were only observed once. Four UK lineages (UK244, UK606, UK1049, UK1162) were observed three times each, and only in key workers and their families. As these lineages were not observed in hospitals or care facilities, it is likely that these key workers became infected in the community. It also indicates that these key workers were unlikely to have passed the virus to patients, and providing evidence that infection control measures were effective in these cases. All UK lineages observed in household members of key workers were also seen in the key workers. Definitive confirmation of transmission, and the direction of transmission, cannot be inferred from the genome sequences due to the low evolutionary rate of the virus, which we have observed in our dataset as approximately two changes per month. Over time most lineages became dormant or extinct, with some expanding rapidly such as UK448 and then disappearing just as quickly over a three week period. Caution must be exercised when making comparisons across different geographical regions, as the UK accounts for 64% (39483 out of 61740 as at 2020-09-01) of all publicly sequenced SARS-CoV-2 genomes. Out of 103 countries that have made SARS-CoV-2 genomes publicly available through GISAID, only the UK, Australia, Spain, India and the USA have sequenced more genomes than have been sequenced in Norfolk alone for this paper.

## Conclusion

We provide an in-depth examination of the genomic epidemiology of SARS-CoV-2 within a single geographical region covering the whole of the first wave of the pandemic from March to August 2020. We sequenced the genomes from 172 SARS-CoV-2-positive samples per 100,000 population (1,035 cases), representing 42.6% of all positive samples collected through the Microbiology Department at NNUH. From this, we identified 100 distinct lineages in the region, corresponding to multiple parallel introductions of the virus (n = >26).

Dense sequencing of the virus provided actionable information for pandemic management including: identifying a sublineage associated with care facilities, ruling out a large nosocomial outbreak in a hospital, showing no evidence of reinfection in longitudinal samples, and confirming an outbreak at a food processing facility while allowing for spillover into the community to be monitored. These achievements were only possible through the collaborative efforts of scientists (data and molecular), clinicians, data managers and epidemiologists.The large-scale genome sequencing of SARS-CoV-2-positive samples has provided valuable additional data for public health epidemiology in the Norfolk region, and will continue to help identify and untangle hidden transmission chains as the pandemic evolves.

## Data Availability

Consensus sequences were deposited in GISAID if they met its minimum QC threshold. Raw sequence data were deposited in the European Nucleotide Archive (ENA). Accession numbers detailed in Supplementary Tables 1-3.

## Ethical approval

The COVID-19 Genomics UK Consortium has been given approval by Public Health England’s Research Ethics and Governance Group (PHE R&D Ref: NR0195).

## Acknowledgements

Thanks to the COG-UK Consortium Study Group for their contributions. Thank you to Dr Judith Pell for her help and insightful comments on this manuscript. We gratefully acknowledge the submitters to GISAID, full details listed in Supplementary Material 2.

## Financial declaration

LG received a partial support for his PhD from Roche.

## Author contributions

All authors have read this manuscript and consented to its publication.

The study was designed and conceived by JOG, AJP. Paper writing was by AJP, AEM. Metadata analysis was by AJP, TLV, AEM, EM. Bioinformatics analysis and informatics were performed by TLV, LOM, NFA, AJP. Sequencing and library preparation was performed by DJB, SR, GLK, AA, APT, AB, AJT, NMT, RG, JOG. Clinical diagnostics and extractions were managed by AK, SD, RP, NE, EM, LC, LB, RD. DA, CB undertook discharge analysis of care facility residents. Genomic epidemiological analyses performed by AJP with oversight from MC, LS, SD, EM, LB. Samples and metadata were collected by LG, WP, RD, AB, AVG, EMA, AK, TS, AJT and MD and biobanked by RS, RNA was extracted by AB, AJT. Risk assessments were by GLK, RAK, JW. Project management and oversight was by GLK, JOG, AJP, LM, MW, AEM, JW. Funding for the project was secured by JOG, AJP.

## Supplementary Figures

**Supplementary Figure 1:**
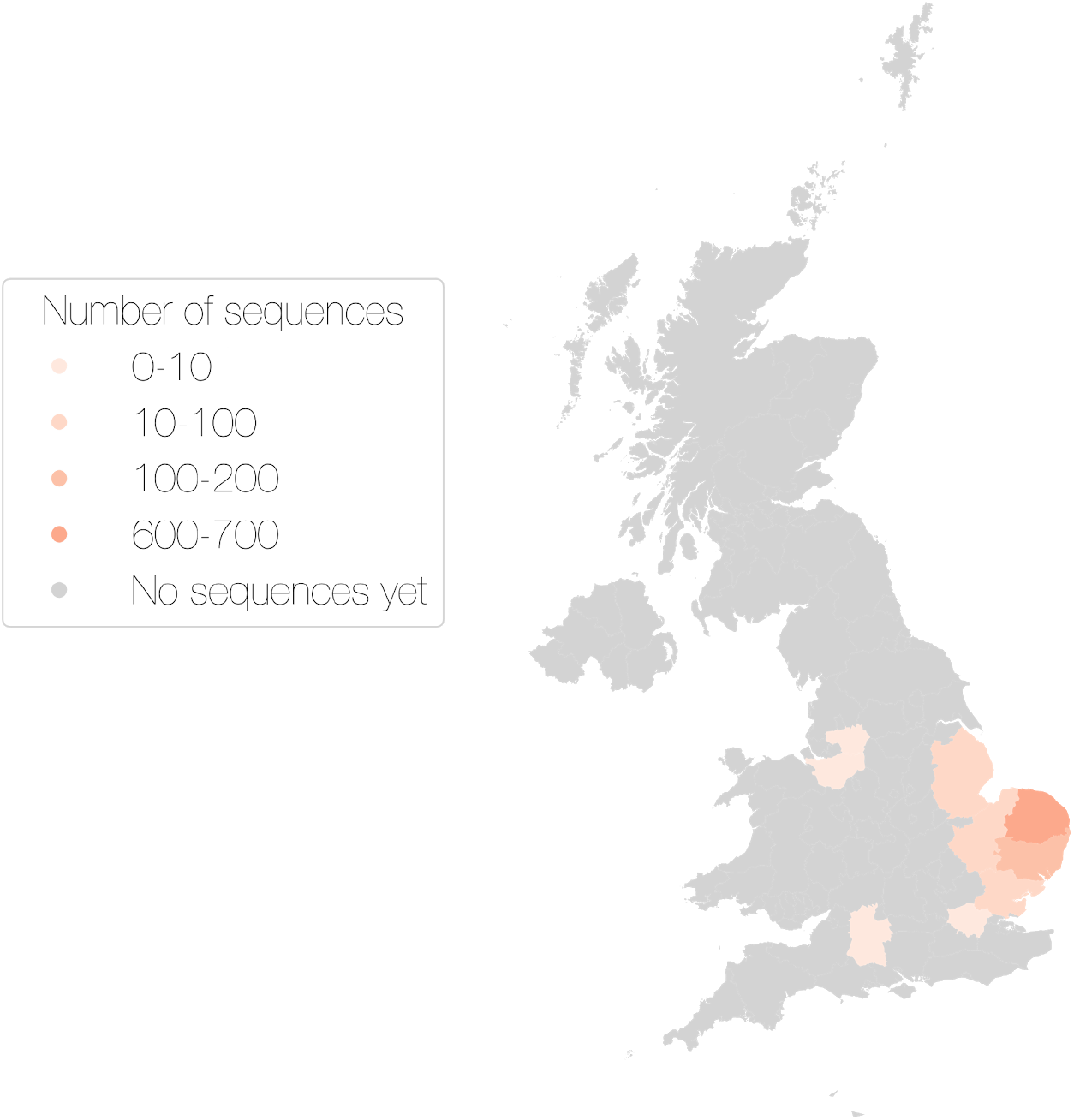
The reported home address of the cases sampled, anonymised to region level.

**Supplementary Figure 2:**
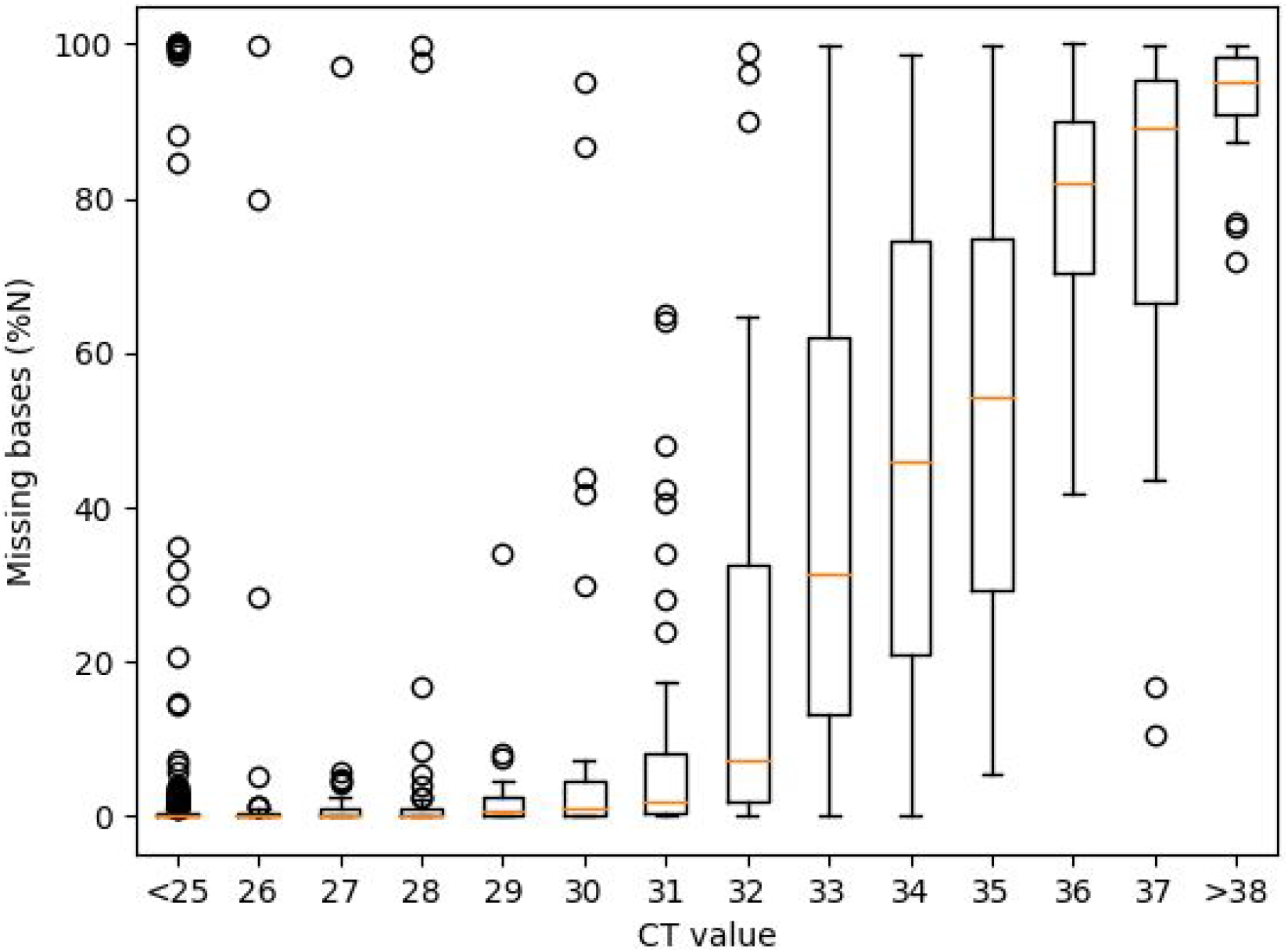
The percentage of missing bases in each consensus genome against the Ct value, where 0 means the consensus genome has no missing bases compared to the Wuhan Hu-1 reference genome (accession MN908947.3) and 100 means the consensus genome is missing every base.

**Supplementary Figure 3:**
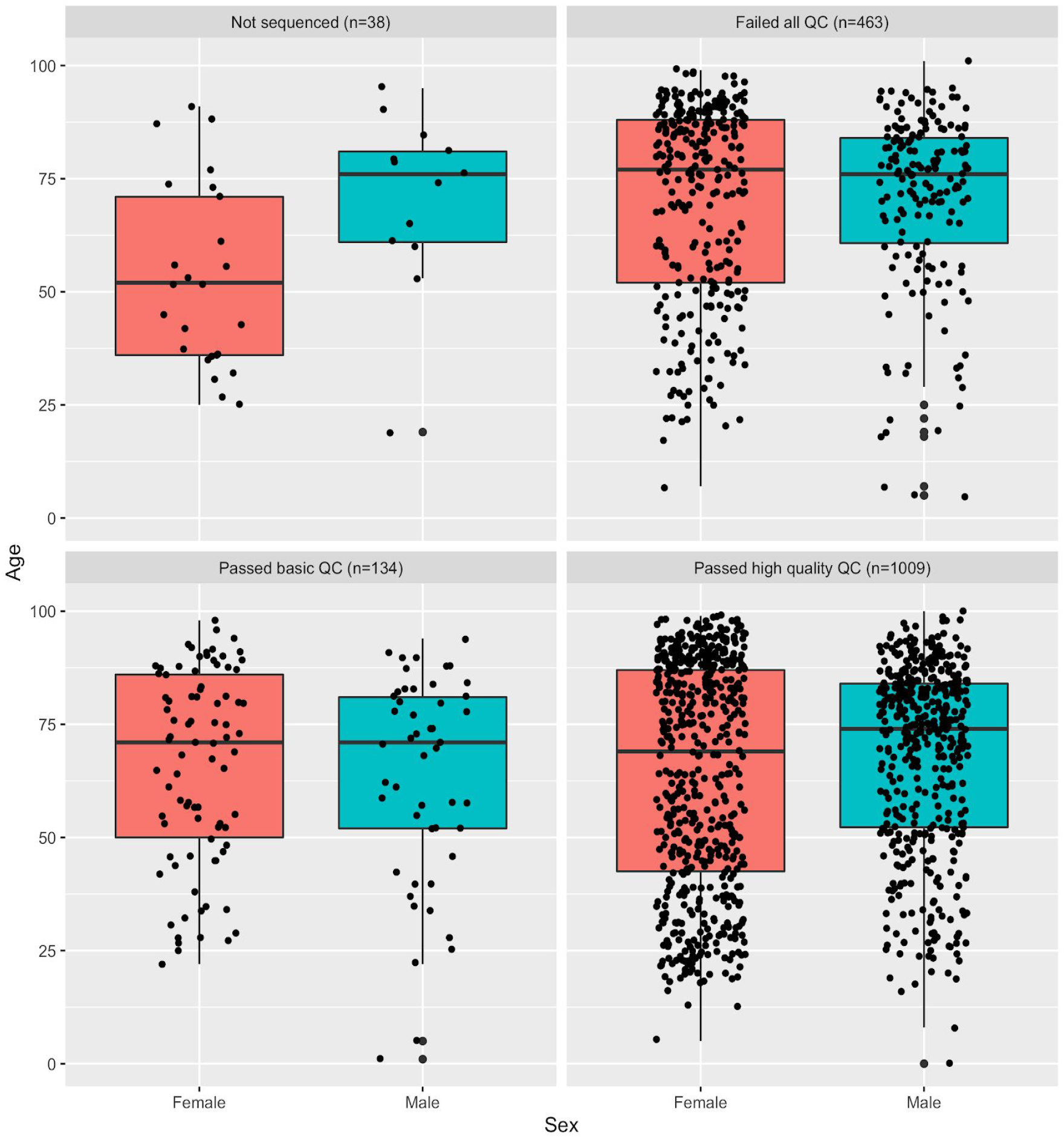
The samples, for which age and sex information were available, categorised by age, sex of the SARS-CoV-2 positive individual and viral genome sequencing quality.

**Supplementary Figure 4:**
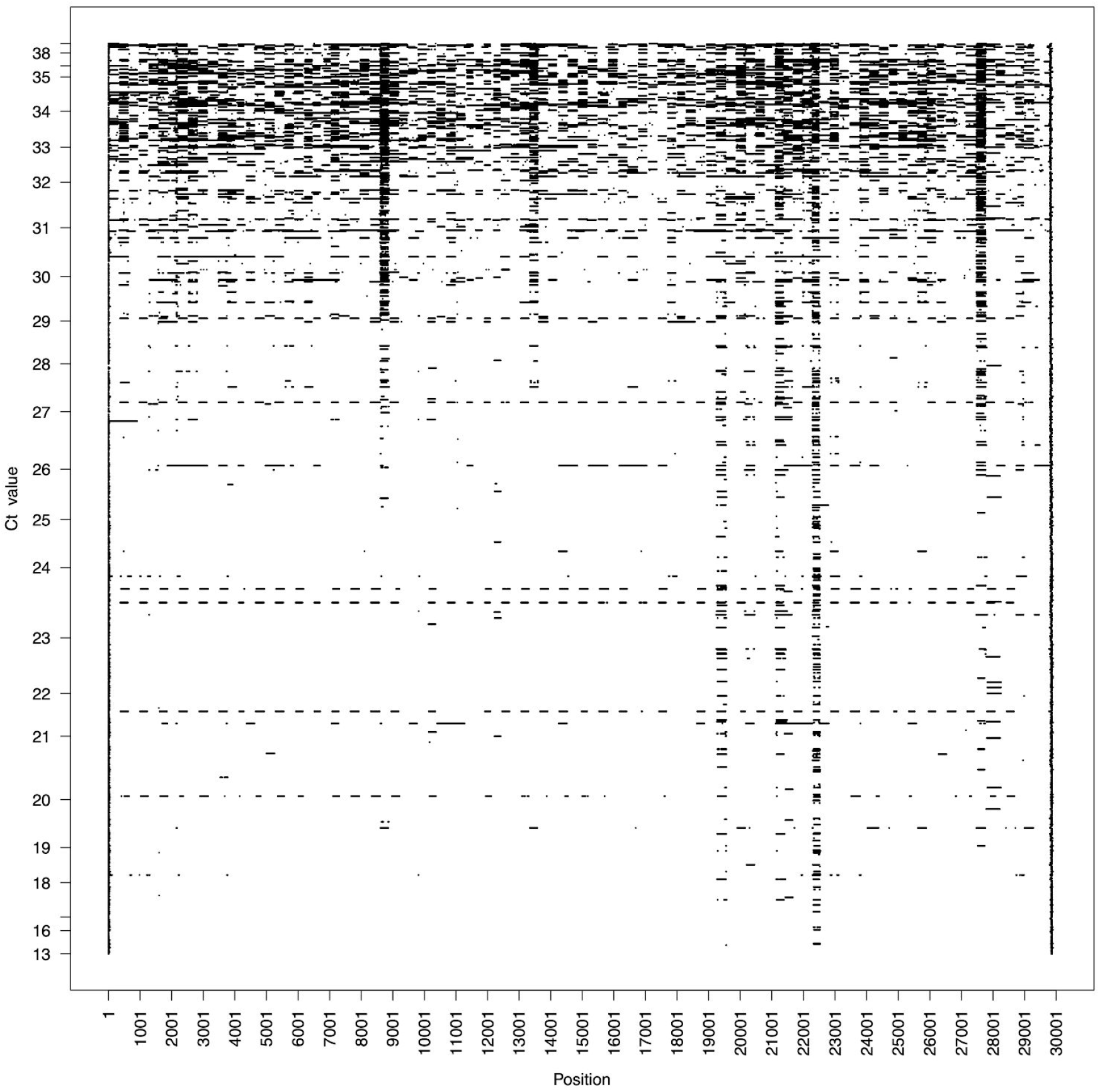
The consensus genomes samples against the Wuhan Hu-1 reference genome (accession MN908947.3), where a black line indicates the absence of data (N) in a consensus genome and white indicates the presence of data.

**Supplementary Figure 5:**
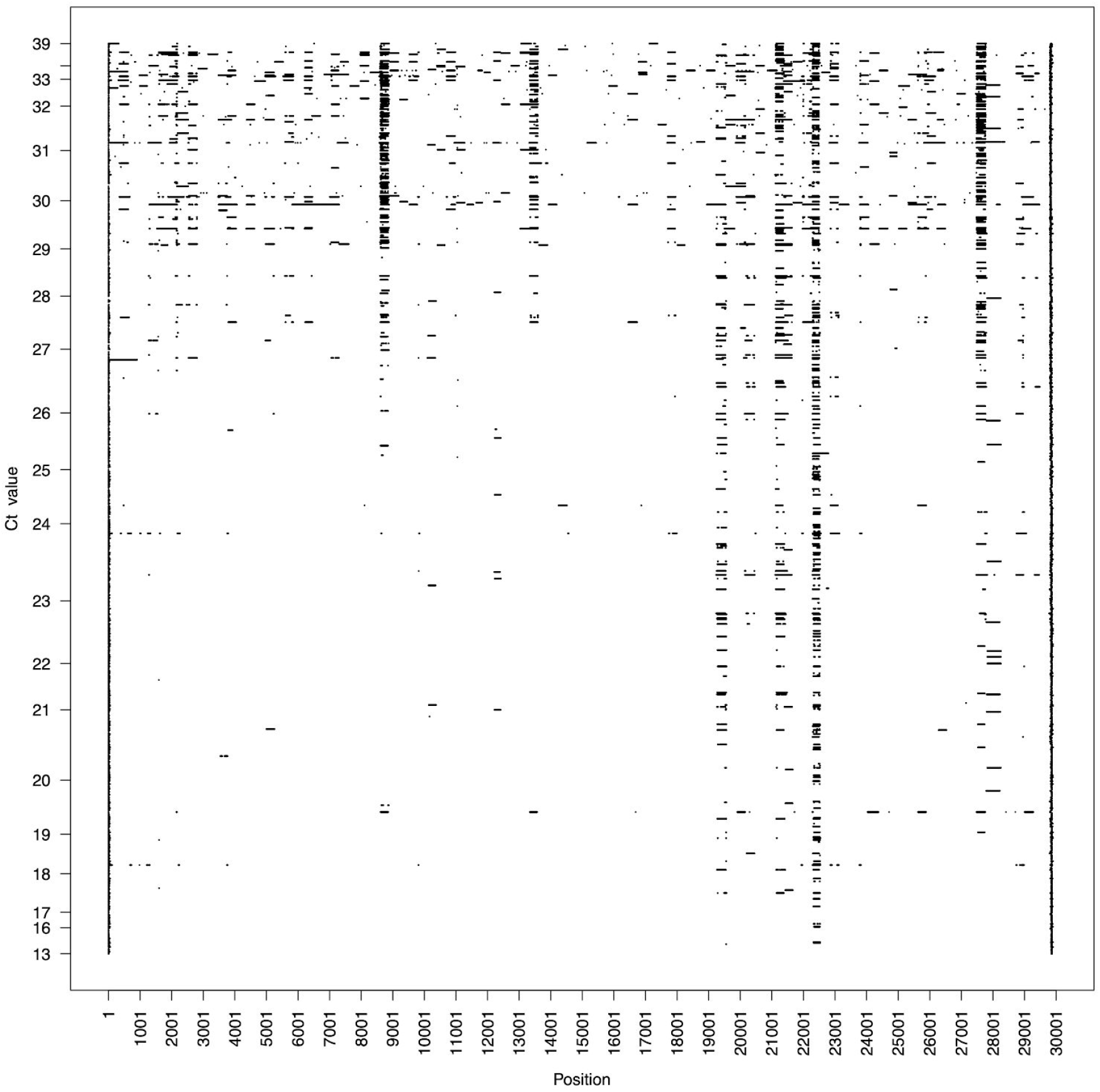
The consensus genomes of all samples passing GISIAD QC against the Wuhan Hu-1 reference genome, where a black line indicates the absence of data (N) in a consensus genome.

**Supplementary Figure 6:**
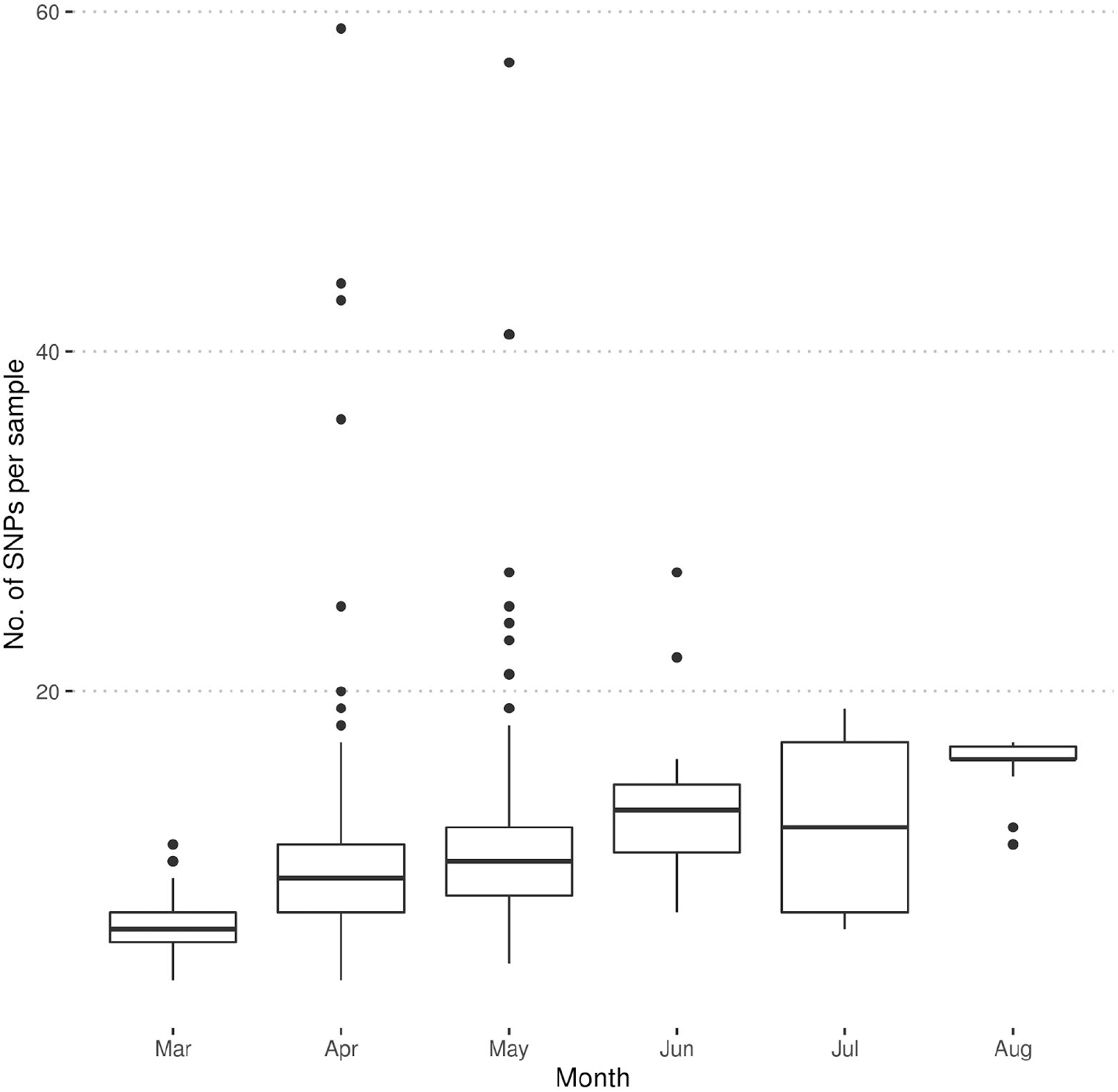
The number of SNPs per sample per month of collection compared to the Wuhan Hu-1 reference genome.

**Supplementary Figure 7:**
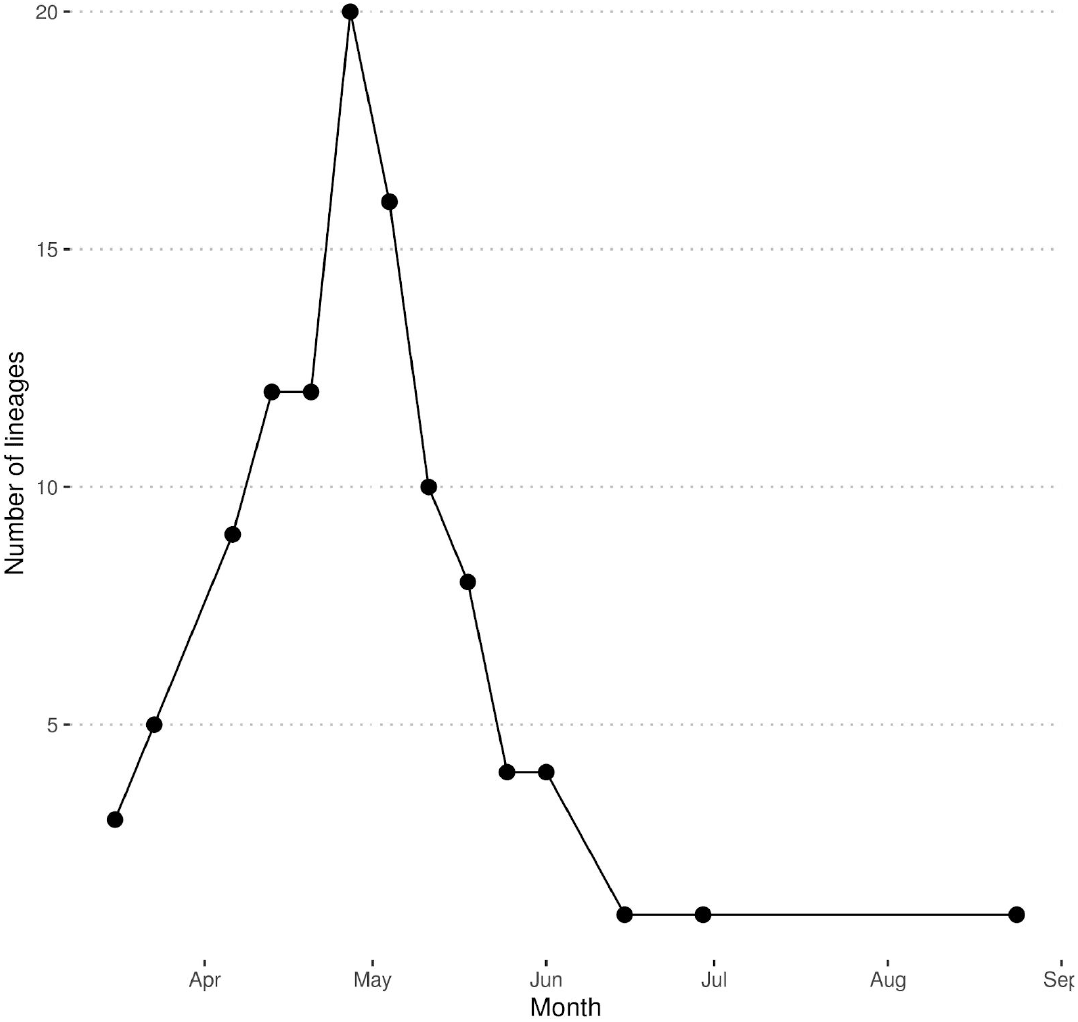
The number of UK lineages observed in a given week in Norfolk.

**Supplementary Figure 8:**
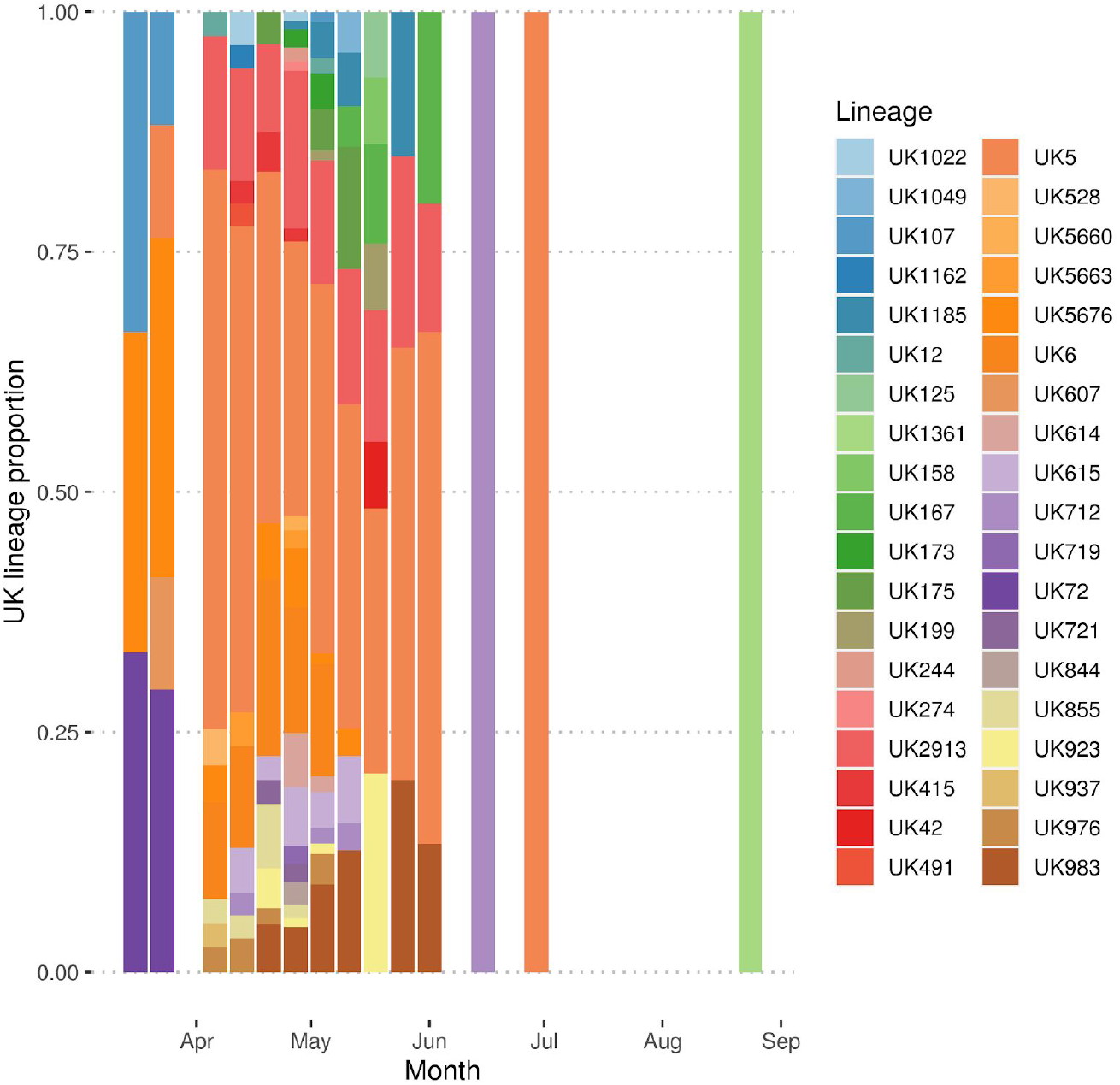
The proportion of samples for each UK lineage identified in Norfolk where the lineage contained two or more representatives.

**Supplementary Figure 9:**
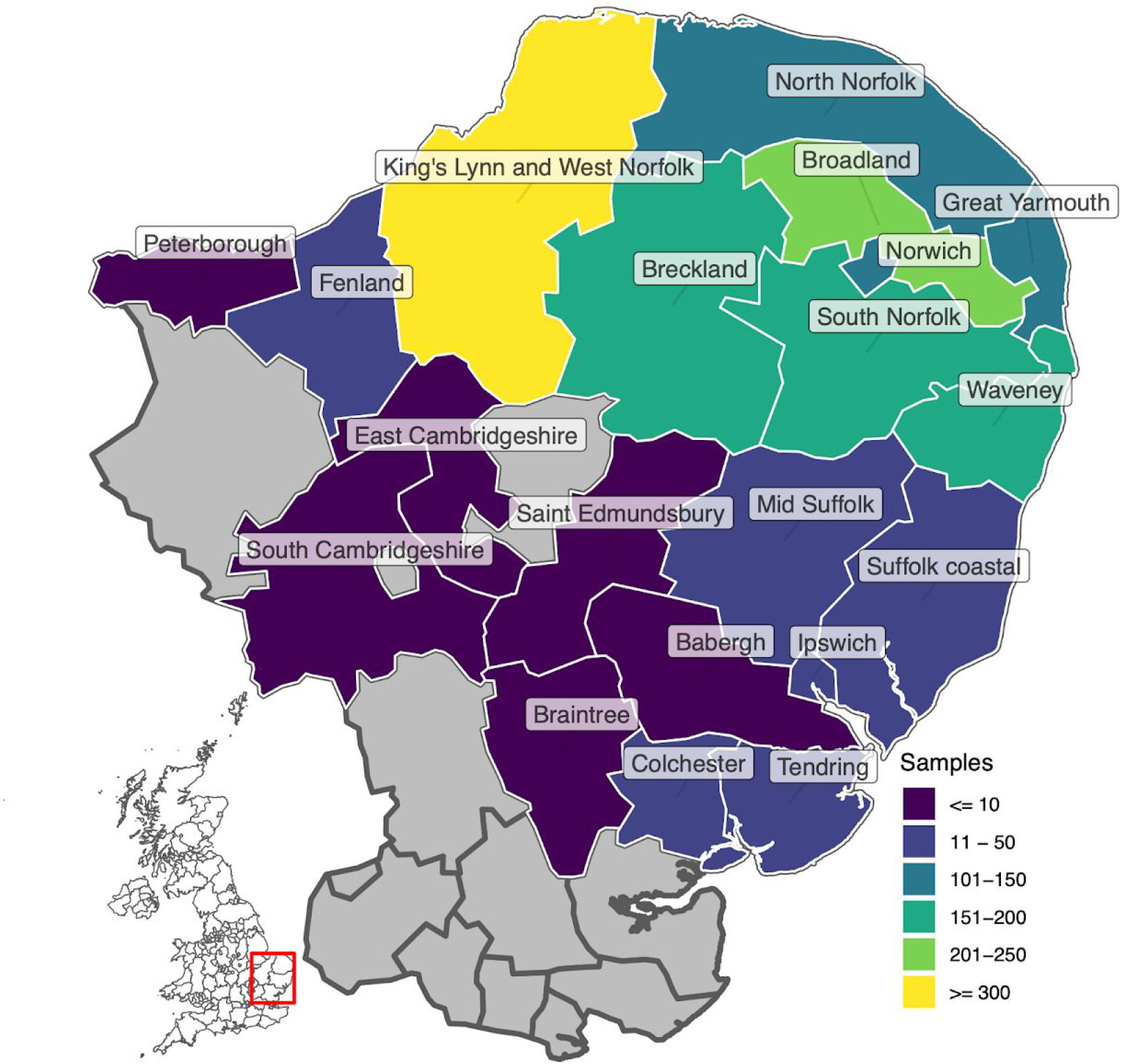
Number of cases sequenced per locality (NUTS level 3).

## Supplementary Material 1

COVID-19 Genomics UK (COG-UK) consortium names and affiliations.

## Supplementary Material 2

GISAID acknowledgement

