## Supplementary Material 1 for "Large scale sequencing of SARS-CoV-2 genomes from one region allows detailed epidemiology and enables local outbreak management"

**Funding acquisition, leadership, supervision, metadata curation, project administration, samples, logistics, Sequencing, analysis, and Software and analysis tools:**
Dr Thomas R Connor PhD^33, 34^	, and Professor Nicholas J Loman PhD^15^.

**Leadership, supervision, sequencing, analysis, funding acquisition, metadata curation, project administration, samples, logistics, and visualisation:**

Dr Samuel C Robson Ph.D ^68^.

**Leadership, supervision, project administration, visualisation, samples, logistics, metadata curation and software and analysis tools:**

Dr Tanya Golubchik PhD ^27^.

**Leadership, supervision, metadata curation, project administration, samples, logistics sequencing and analysis:**

Dr M. Estee Torok FRCP ^8, 10^.

**Project administration, metadata curation, samples, logistics, sequencing, analysis, and software and analysis tools:**

Dr William L Hamilton PhD ^8, 10^.

**Leadership, supervision, samples logistics, project administration, funding acquisition sequencing and analysis:**

Dr David Bonsall PhD	^27^.

**Leadership and supervision, sequencing, analysis, funding acquisition, visualisation and software and analysis tools:**

Dr Ali R Awan	PhD ^74^.

**Leadership and supervision, funding acquisition, sequencing, analysis, metadata curation, samples and logistics:**

Dr Sally Corden PhD ^33^ .

**Leadership supervision, sequencing analysis, samples, logistics, and metadata curation:** Professor Ian Goodfellow PhD ^11^.

**Leadership, supervision, sequencing, analysis, samples, logistics, and Project administration:**

Professor Darren L Smith PhD ^60, 61^.

**Project administration, metadata curation, samples, logistics, sequencing and analysis:**
Dr Martin D Curran PhD ^14^, and Dr Surendra Parmar PhD ^14^**.**

**Samples, logistics, metadata curation, project administration sequencing and analysis:**
Dr James G Shepherd MBChB MRCP	^21^.

**Sequencing, analysis,	project administration, metadata curation and software and analysis tools:**

Dr Matthew D Parker PhD ^38^.

**Leadership, supervision, funding acquisition, samples, logistics, and metadata curation:**
Dr Catherine Moore^33^	.

**Leadership, supervision, metadata curation, samples, logistics, sequencing and analysis:**
Dr Derek J Fairley PhD ^6, 88^, Professor Matthew W Loose PhD ^54^, and Joanne Watkins MSc ^33^.

**Metadata curation, sequencing, analysis, leadership, supervision and software and analysis tools:**

Dr Matthew Bull PhD^33^ , and Dr Sam Nicholls PhD ^15^	.

**Leadership, supervision, visualisation, sequencing, analysis and software and analysis tools:**

Professor David M Aanensen PhD ^1, 30^.

**Sequencing, analysis, samples, logistics, metadata curation, and visualisation:**
Dr Sharon Glaysher ^70^	.

**Metadata curation, sequencing, analysis, visualisation, software and analysis tools:**
Dr Matthew Bashton PhD ^60^, and Dr Nicole Pacchiarini PhD ^33^.

**Sequencing, analysis, visualisation, metadata curation, and software and analysis tools**:
Dr Anthony P Underwood PhD ^1, 30^.

**Funding acquisition, leadership, supervision and project administration:**
Dr Thushan I de Silva PhD ^38^, and Dr Dennis Wang PhD ^38^**.**

**Project administration, samples, logistics, leadership and supervision**:

Dr Monique Andersson PhD ^28^ , Professor Anoop J Chauhan ^70^, Dr Mariateresa de Cesare PhD^26^, Dr Catherine Ludden ^1,3^ , and Dr Tabitha W Mahungu FRCPath ^91^.

**Sequencing, analysis, project administration and metadata curation:**
Dr Rebecca Dewar PhD ^20^, and Martin P McHugh MSc^20^.

**Samples, logistics, metadata curation and project administration:**
Dr Natasha G Jesudason MBChB MRCP FRCPath ^21^, Dr Kathy K Li MBBCh FRCPath	^21^, Dr Rajiv N Shah	BMBS MRCP MSc ^21^, and Dr Yusri Taha MD, PhD ^66^.

**Leadership, supervision, funding acquisition and metadata curation:**
Dr Kate E Templeton PhD ^20^**.**

**Leadership, supervision, funding acquisition, sequencing and analysis:**
Dr Simon Cottrell PhD ^33^, Dr Justin O’Grady PhD ^51^, Professor Andrew Rambaut DPhil ^19^, and Professor Colin P Smith PhD^93^.

**Leadership, supervision, metadata curation	, sequencing and analysis:**
Professor Matthew T.G. Holden PhD	^87^, and Professor Emma C Thomson PhD/FRCP ^21^.

**Leadership, supervision, samples, logistics and metadata curation**:
Dr Samuel Moses MD ^81, 82^.

**Sequencing, analysis, leadership, supervision, samples and logistics:**
Dr Meera Chand ^7^, Dr Chrystala Constantinidou PhD ^71^, Professor Alistair C Darby PhD ^46^, Professor Julian A Hiscox PhD ^46^, Professor Steve Paterson PhD ^46^, and Dr Meera Unnikrishnan PhD ^71^**.**

**Sequencing, analysis, leadership and supervision and software and analysis tools:**
Dr Andrew J Page PhD ^51^, and Dr Erik M Volz PhD^96^.

**Samples, logistics, sequencing, analysis and metadata curation:**
Dr Charlotte J Houldcroft PhD ^8^, Dr Aminu S Jahun PhD ^11^, Dr James P McKenna PhD ^88^, Dr Luke W Meredith PhD ^11^, Dr Andrew Nelson PhD ^61^, Sarojini Pandey MSc ^72^, and Dr Gregory R Young PhD ^60^.

**Sequencing, analysis, metadata curation, and software and analysis tools:**
Dr Anna Price PhD^34^, Dr Sara Rey PhD ^33^, Dr Sunando Roy PhD ^41^, Dr Ben Temperton Ph.D ^49^, and Matthew Wyles ^38^.

**Sequencing, analysis, metadata curation and visualisation:**

Stefan	Rooke	MSc^19^,	and Dr Sharif Shaaban PhD^87^.

**Visualisation, sequencing, analysis and software and analysis tools:**
Dr Helen Adams PhD ^35^, Dr Yann Bourgeois Ph.D ^69^, Dr Katie F Loveson Ph.D ^68^, Áine O'Toole MSc^19^, and Richard Stark MSc ^71^.

**Project administration, leadership and supervision:**

Dr Ewan M Harrison PhD ^1, 3^, David Heyburn ^33^, and Professor Sharon J Peacock ^2, 3^

**Project administration and funding acquisition:**

Dr David Buck PhD^26^ , and Michaela John BSc Hons ^36^

**Sequencing, analysis and project administration:**

Dorota	Jamrozy ^1^, and Dr Joshua Quick PhD ^15^

**Samples, logistics, and project administration:**

Dr Rahul Batra MD^78^, Katherine L Bellis BSc (Hons) ^1, 3^, Beth Blane BSc ^3^ , Sophia T Girgis MSc ^3^, Dr Angie Green PhD ^26^, Anita Justice MSc ^28^ , Dr Mark Kristiansen PhD ^41^ , and Dr Rachel J Williams PhD ^41^.

**Project administration, software and analysis tools:**

Radoslaw Poplawski BSc ^15^.

**Project administration and visualisation:**

Dr Garry P Scarlett Ph.D^69^.

**Leadership, supervision, and funding acquisition:**

Professor John A Todd PhD ^26^	, Dr Christophe Fraser	PhD ^27^,	Professor Judith Breuer MD ^40,41^, Professor Sergi Castellano PhD ^41^, Dr Stephen L Michell PhD ^49^, Professor Dimitris Gramatopoulos PhD, FRCPath^73^, and Dr Jonathan Edgeworth PhD, FRCPath ^78^.

**Leadership, supervision and metadata curation:**
Dr Gemma L Kay PhD ^51^.

**Leadership, supervision, sequencing and analysis:**
Dr Ana da Silva Filipe PhD ^21^ , Dr Aaron R Jeffries PhD ^49^, Dr Sascha Ott PhD ^71^, Professor Oliver Pybus ^24^ , Professor David L Robertson PhD ^21^, Dr David A Simpson PhD ^6^ , and Dr Chris Williams MB BS^33^.

**Samples, logistics, leadership and supervision:**

Dr Cressida Auckland FRCPath ^50^, Dr John Boyes MBChB^83^, Dr Samir Dervisevic FRCPath^52^ , Professor Sian Ellard FRCPath^49, 50^ , Dr Sonia Goncalves^1^, Dr Emma J Meader FRCPath ^51^, Dr Peter Muir PhD^2^, Dr Husam Osman PhD ^95^, Reenesh Prakash MPH^52^, Dr Venkat Sivaprakasam PhD^18^, and Dr Ian B Vipond PhD^2^.

**Leadership, supervision and visualisation**

Dr Jane AH Masoli MBChB ^49, 50^.

**Sequencing, analysis and metadata curation**

Dr Nabil-Fareed Alikhan PhD ^51^, Matthew Carlile BSc ^54^, Dr Noel Craine DPhil ^33^, Dr Sam T Haldenby PhD ^46^, Dr Nadine Holmes PhD ^54^, Professor Ronan A Lyons MD ^37^, Dr Christopher Moore PhD ^54^, Malorie Perry MSc ^33^ , Dr Ben Warne	MRCP^80^, and Dr Thomas Williams MD ^19^.

**Samples, logistics and metadata curation:**

Dr Lisa Berry PhD ^72^, Dr Andrew Bosworth PhD ^95^ ,Dr Julianne Rose Brown PhD^40^, Sharon Campbell MSc	^67^, Dr Anna Casey PhD	^17^, Dr Gemma Clark PhD ^56^, Jennifer Collins BSc ^66^, Dr Alison Cox PhD ^43,^ ^44^ , Thomas Davis MSc ^84^, Gary Eltringham BSc ^66^, Dr Cariad Evans ^38, 39^ , Dr Clive Graham MD ^64^, Dr Fenella Halstead PhD ^18^, Dr Kathryn Ann Harris PhD ^40^, Dr Christopher Holmes PhD ^58^, Stephanie Hutchings ^2^ , Professor Miren Iturriza-Gomara PhD ^46^ , Dr Kate Johnson ^38, 39^, Katie Jones MSc ^72^, Dr Alexander J Keeley MRCP ^38^, Dr Bridget A Knight PhD ^49, 50^ , Cherian Koshy MSc, CSci, FIBMS ^90^, Steven Liggett ^63^ , Hannah Lowe MSc ^81^ , Dr Anita O Lucaci PhD ^46^ , Dr Jessica Lynch PhD MBChB ^25, 29^ , Dr Patrick C McClure PhD ^55^ , Dr Nathan Moore MBChB ^31^ , Matilde Mori BSc ^25, 29, 32^ , Dr David G Partridge FRCP, FRCPath ^38, 39^ , Pinglawathee Madona ^43, 44^ , Hannah M Pymont MSc ^2^ , Dr Paul Anthony Randell MBBCh ^43, 44^ , Dr Mohammad Raza ^38, 39^ , Felicity Ryan MSc ^81^ , Dr Robert Shaw FRCPath ^28^, Dr Tim J Sloan PhD ^57^ , and Emma Swindells BSc ^65^ .

**Sequencing, analysis, Samples and logistics:**

Alexander Adams BSc	^33^, Dr Hibo Asad PhD ^33^, Alec Birchley MSc ^33^ , Tony Thomas Brooks BSc (Hons) ^41^, Dr Giselda Bucca PhD ^93^, Ethan Butcher ^70^, Dr Sarah L Caddy PhD ^13^, Dr Laura G Caller PhD ^2, 3, 12^ , Yasmin Chaudhry BSc ^11^, Jason Coombes BSc (HONS) ^33^, Michelle Cronin ^33^, Patricia L Dyal MPhil ^41^, Johnathan M Evans MSc ^33^,Laia Fina ^33^, Bree Gatica-Wilcox MPhil ^33^, Dr Iliana Georgana PhD ^11^, Lauren Gilbert A-Levels ^33^ , Lee Graham BSc ^33^, Danielle C Groves BA ^38^, Grant Hall BSc ^11^, Ember Hilvers MPH	^33^ , Dr Myra Hosmillo PhD ^11^, Hannah Jones ^33^, Sophie Jones MSc ^33^, Fahad	A Khokhar BSc	^13^ , Sara Kumziene-Summerhayes MSc ^33^, George MacIntyre-Cockett BSc ^26^, Dr Rocio T Martinez Nunez PhD ^94^ , Dr Caoimhe McKerr PhD ^33^ , Dr Claire McMurray PhD ^15^,	Dr Richard Myers ^7^, Yasmin Nicole Panchbhaya BSc ^41^ , Malte L Pinckert MPhil ^11^ , Amy Plimmer ^33^ , Dr Joanne Stockton PhD ^15^ , Sarah Taylor ^33^ , Dr Alicia Thornton ^7^ , Amy Trebes MSc ^26^ , Alexander J Trotter MRes ^51^ ,Helena Jane Tutill BSc ^41^ ,Charlotte A Williams BSc ^41^ , Anna Yakovleva BSc ^11^ and Dr Wen C Yew PhD ^62^.

**Sequencing, analysis and software and analysis tools:**
Dr Mohammad T Alam PhD ^71^	, Dr Laura Baxter PhD ^71^, Olivia Boyd MSc ^96^ , Dr Fabricia F. Nascimento PhD ^96^, Timothy M Freeman MPhil ^38^, Lily Geidelberg MSc ^96^, Dr Joseph Hughes PhD ^21^, David Jorgensen MSc ^96^, Dr Benjamin B Lindsey MRCP ^38^, Dr Richard J Orton PhD ^21^ , Dr Manon Ragonnet-Cronin PhD ^96^ Joel Southgate MSc ^33, 34,^ and Dr Sreenu Vattipally PhD ^21^.

**Samples, logistics and software and analysis tools:**

Dr Igor	Starinskij MSc MRCP ^23^.

**Visualisation and software and analysis tools:**
Dr Joshua B Singer PhD ^21^ , Dr Khalil Abudahab PhD ^1, 30^, Leonardo de Oliveira Martins PhD^51^ , Dr Thanh Le-Viet PhD	^51^ ,Mirko Menegazzo ^30^ ,Ben EW Taylor Meng ^1, 30^, and Dr Corin A Yeats PhD ^30^.

**Project Administration:**

Sophie Palmer	^3^, Carol M Churcher ^3^ , Dr Alisha Davies ^33^, Elen De Lacy MSc ^33^, Fatima Downing ^33^, Sue Edwards ^33^ , Dr Nikki Smith	PhD ^38^ , and Dr Frances Bolt PhD ^44, 45^	.

**Leadership and supervision:**

Dr. Alex Alderton ^1^, Dr Matt Berriman ^1^, Ian G Charles ^51^, Dr Nicholas Cortes MBChB ^31^ ,Dr Tanya Curran PhD ^88^ , Prof John Danesh ^1^, Dr Sahar Eldirdiri MBBS, MSC FRCPath ^84^, Dr Ngozi Elumogo FRCPath ^52^, Prof Andrew Hattersley FRS ^49, 50^, Professor Alison Holmes MD ^44, 45^, Dr Robin Howe ^33^, Dr Rachel Jones ^33^ , Anita Kenyon MSc ^84^, Prof Robert A Kingsley PhD ^51^, Professor Dominic Kwiatkowski ^1, 9^, Dr Cordelia Langford^1^, Dr Jenifer Mason MBBS ^48^, Dr Alison E Mather PhD ^51^, Lizzie Meadows MA ^51^, Dr Sian Morgan FRCPath ^36^, Dr James Price PhD ^44, 45^, Trevor I Robinson MSc ^48^ , Dr Giri Shankar ^33^ , John Wain ^51^, and Dr Mark A Webber PhD^51^	.

**Metadata curation:**

Dr Declan T Bradley PhD ^5, 6^ ,Dr Michael R Chapman PhD ^1, 3, 4^ , Dr Derrick Crooke ^28^ , Dr David Eyre PhD ^28^, Professor Martyn Guest PhD^34^ , Huw Gulliver ^34^ , Dr Sarah Hoosdally ^28^ , Dr Christine Kitchen PhD ^34^ , Dr Ian Merrick PhD ^34^, Siddharth Mookerjee MPH ^44, 45^ , Robert Munn	BSc ^34^ , Professor Timothy Peto PhD^28^, Will Potter ^52^, Dr Dheeraj K Sethi MBBS ^52^, Wendy Smith ^56^ ,Dr Luke B Snell MB BS ^75, 94^ , Dr Rachael Stanley PhD ^52^ , Claire Stuart ^52^ and Dr Elizabeth Wastenge MD ^20^.

**Sequencing and analysis:**

Dr Erwan Acheson PhD ^6^ , Safiah Afifi BSc ^36^ , Dr Elias Allara	MD PhD ^2, 3^ , Dr Roberto Amato ^1^, Dr Adrienn Angyal PhD^38^, Dr Elihu Aranday-Cortes PhD/DVM^21^ , Cristina Ariani ^1^, Jordan Ashworth ^19^,	Dr Stephen Attwood ^24^, Alp Aydin MSci ^51^ , David J Baker BEng ^51^ , Dr Carlos E Balcazar PhD ^19^, Angela Beckett MSc	^68^ Robert Beer BSc ^36^ ,	Dr Gilberto Betancor	PhD^76^, Emma	Betteridge ^1^ , Dr David Bibby ^7^ , Dr Daniel Bradshaw	^7^ , Catherine Bresner Bsc(Hons) ^34^, Dr Hannah E Bridgewater PhD ^71^ , Alice Broos BSc (Hons) ^21^ , Dr Rebecca Brown PhD ^38^ , Dr Paul E Brown PhD ^71^, Dr Kirstyn Brunker PhD ^22^ , Dr Stephen N Carmichael PhD ^21^ , Jeffrey K. J. Cheng MSc ^71^, Dr Rachel Colquhoun DPhil ^19^ , Dr Gavin Dabrera ^7^ , Dr Johnny Debebe PhD ^54^, Eleanor Drury ^1^, Dr Louis du Plessis ^24^ , Richard Eccles MSc ^46^, Dr Nicholas Ellaby ^7^, Audrey Farbos MSc ^49^, Ben Farr ^1^ ,Dr Jacqueline Findlay PhD ^41^ , Chloe L Fisher MSc ^74^, Leysa Marie Forrest MSc ^41^, Dr Sarah Francois ^24^, Lucy R. Frost BSc ^71^, William Fuller BSc ^34^ , Dr Eileen Gallagher ^7^ , Dr Michael D Gallagher PhD^19^ , Matthew Gemmell MSc ^46^, Dr Rachel AJ Gilroy PhD ^51^, Scott Goodwin ^1^, Dr Luke R Green PhD ^38^ ,Dr Richard Gregory PhD ^46^ ,Dr Natalie Groves ^7^ ,Dr James W Harrison PhD ^49^, Hassan Hartman ^7^ , Dr Andrew R Hesketh PhD^93^,Verity Hill ^19^, Dr Jonathan Hubb ^7^ ,Dr Margaret Hughes PhD^46^ ,Dr David K Jackson ^1^ ,Dr Ben Jackson PhD ^19^ ,Dr Keith James ^1^ ,Natasha Johnson BSc (Hons)^21^ ,Ian Johnston ^1^, Jon-Paul Keatley ^1^, Dr Moritz Kraemer ^24^, Dr Angie Lackenby ^7^, Dr Mara Lawniczak ^1^ , Dr David Lee ^7^, Rich Livett ^1^, Stephanie Lo ^1^, Daniel Mair BSc (Hons) ^21^, Joshua Maksimovic FD sport science ^36^, Nikos Manesis ^7^ ,Dr Robin Manley Ph.D ^49^, Dr Carmen Manso ^7^ ,Dr Angela Marchbank BSc ^34^ ,Dr Inigo Martincorena ^1^ ,Dr Tamyo Mbisa ^7^, Kathryn McCluggage MSC ^36^,Dr JT McCrone PhD ^19^, Shahjahan Miah ^7^ , Michelle L Michelsen BSc ^49^ , Dr Mari Morgan PhD ^33^, Dr Gaia Nebbia PhD, FRCPath ^78^,Charlotte Nelson MSc ^46^ ,Jenna Nichols BSc (Hons) ^21^ ,Dr Paola Niola PhD ^41^ ,Dr Kyriaki Nomikou PhD^21^ ,Steve Palmer ^1^ , Dr. Naomi Park ^1^, Dr Yasmin A Parr PhD^21^ ,Dr Paul J Parsons PhD ^38^ , Vineet Patel ^7^ ,Dr. Minal Patel ^1^ ,Clare Pearson MSc ^2, 1^ ,Dr Steven Platt ^7^ ,Christoph Puethe ^1^, Dr. Mike Quail ^1^,Dr JaynaRaghwani ^24^ , Dr Lucille Rainbow PhD ^46^ ,Shavanthi Rajatileka ^1^ ,Dr Mary Ramsay ^7^ ,Dr Paola C Resende Silva PhD ^41, 42^, Steven Rudder 51, Dr Chris Ruis ^3^ ,Dr Christine M Sambles PhD ^49^ ,Dr Fei Sang PhD ^54^ ,Dr Ulf Schaefer^7^ ,Dr Emily Scher PhD ^19^ ,Dr. Carol Scott ^1^ ,Lesley Shirley ^1^, Adrian W Signell BSc ^76^ ,John Sillitoe ^1^ ,Christen Smith ^1^ ,Dr Katherine L Smollett PhD ^21^ ,Karla Spellman FD ^36^ ,Thomas D Stanton BSc ^19^ ,Dr David J Studholme PhD ^49^ ,Ms Grace Taylor-Joyce BSc ^71^ ,Dr Ana P Tedim PhD ^51^ ,Dr Thomas Thompson PhD^6^ ,Dr Nicholas M Thomson PhD ^51^ ,Scott Thurston^1^ ,Lily Tong PhD ^21^ ,Gerry Tonkin-Hill ^1^, Rachel M Tucker MSc ^38^ , Dr Edith E Vamos PhD ^4^,Dr Tetyana Vasylyeva^24^ , Joanna Warwick-Dugdale BSc ^49^ , Danni Weldon ^1^ ,Dr Mark Whitehead PhD ^46^ ,Dr David Williams ^7^,Dr Kathleen A Williamson PhD^19^,Harry D Wilson BSc ^76^,Trudy Workman HNC ^34^ ,Dr Muhammad Yasir PhD ^51^, Dr Xiaoyu Yu PhD ^19^, and Dr Alex Zarebski ^24^.

**Samples and logistics:**

Dr Evelien M Adriaenssens PhD ^51^, Dr Shazaad S Y Ahmad MSc ^2, 47^ , Adela Alcolea-Medina MPharm ^59, 77^ ,Dr John Allan PhD^60^, Dr Patawee Asamaphan PhD^21^, Laura Atkinson MSc ^40^, Paul Baker MD ^63^, Professor Jonathan Ball PhD ^55^, Dr Edward Barton MD^64^, Dr. Mathew A Beale^1^, Dr. Charlotte Beaver^1^ , Dr Andrew Beggs PhD^16^, Dr Andrew Bell PhD^51^, Duncan J Berger ^1^, Dr Louise Berry. ^56^, Claire M Bewshea MSc ^49^, Kelly Bicknell ^70^, Paul Bird ^58^, Dr Chloe Bishop ^7^ , Dr Tim Boswell ^56^, Cassie Breen BSc^48^, Dr Sarah K Buddenborg^1^, Dr Shirelle Burton-Fanning MD ^66,^ Dr Vicki Chalker ^7^, Dr Joseph G Chappell PhD ^55^, Themoula Charalampous MSc ^78, 94^, Claire Cormie^3^, Dr Nick Cortes PhD^29, 25^, Dr Lindsay J Coupland PhD ^52^, Angela Cowell MSc^48^ , Dr Rose K Davidson PhD ^53 ,^ Joana Dias MSc^3^ , Dr Maria Diaz PhD^51^ , Thomas Dibling^1^, Matthew J Dorman^1^, Dr Nichola Duckworth^57^, Scott Elliott^70^, Sarah Essex^63^, Karlie Fallon ^58^ , Theresa Feltwell ^8^ , Dr Vicki M Fleming PhD ^56^, Sally Forrest BSc ^3^, Luke Foulser^1^, Maria V Garcia-Casado^1^, Dr Artemis Gavriil PhD ^41^, Dr Ryan P George PhD^47^, Laura Gifford MSc ^33^, Harmeet K Gill PhD^3^, Jane Greenaway MSc^65^, Luke Griffith Bsc^53^, Ana Victoria Gutierrez^51^, Dr Antony D Hale MBBS^85^, Dr Tanzina Haque FRCPath, PhD^91^, Katherine L Harper MBiol^85^, Dr Ian Harrison ^7^ , Dr Judith Heaney PhD^89^, Thomas Helmer ^58^, Ellen E Higginson PhD ^3^ , Richard Hopes ^2^, Dr Hannah C Howson-Wells PhD ^56^, Dr Adam D Hunter ^1^, Robert Impey ^70^, Dr Dianne Irish-Tavares FRCPath ^91^, David A Jackson^1^ , Kathryn A Jackson MSc ^46^ , Dr Amelia Joseph ^56^, Leanne Kane ^1^, Sally Kay ^1^, Leanne M Kermack MSc ^3^, Manjinder Khakh ^56^, Dr Stephen P Kidd PhD^29, 25,31^, , Dr Anastasia Kolyva PhD ^51^, Jack CD Lee BSc ^40^, Laura Letchford ^1^ , Nick Levene MSc^79^, Dr LisaJ Levett PhD ^89^, Dr Michelle M Lister PhD ^56^, Allyson Lloyd ^70^ , Dr Joshua Loh PhD^60^ , Dr Louissa R Macfarlane-Smith PhD^85^, Dr Nicholas W Machin MSc ^2 , 47^, Mailis Maes M.phil^3^, Dr Samantha McGuigan ^1^, Liz McMinn ^1^, Dr Lamia Mestek-Boukhibar D.Phil ^41^, Dr Zoltan Molnar PhD ^6^, Lynn Monaghan ^79^, Dr Catrin Moore ^27^, Plamena Naydenova BSc ^3^, Alexandra S Neaverson ^1^, Dr. Rachel Nelson PhD ^1^, Marc O Niebel MSc^21^ , Elaine O'Toole BSc ^48^ , Debra Padgett BSc ^64^, Gaurang Patel ^1^ , Dr Brendan AI Payne MD ^66^, Liam Prestwood ^1^, Dr Veena Raviprakash MD^67^, Nicola Reynolds PhD^86^ Dr Alex Richter PhD ^16^, Dr Esther Robinson PhD^95^, Dr Hazel A Rogers^1^, Dr Aileen Rowan PhD ^96^, Garren Scott BSc ^64^, Dr Divya Shah PhD^40^, Nicola Sheriff BSc ^67^, Dr Graciela Sluga MD - MSc^92^ , Emily Souster^1^, Dr. Michael Spencer-Chapman^1^, Sushmita Sridhar BSc^1, 3^, Tracey Swingler ^53^, Dr Julian Tang^58^, Professor Graham P Taylor DSc^96^, Dr Theocharis Tsoleridis PhD^55^, Dr Lance Turtle PhD MRCP^46^, Dr Sarah Walsh ^57^, Dr Michelle Wantoch PhD ^86^, Joanne Watts BSc^48^ , Dr Sheila Waugh MD^66^, Sam Weeks^41^, Dr Rebecca Williams BMBS ^31^, Dr Iona Willingham^56^, Dr Emma L Wise PhD ^25, 29, 31^, Victoria Wright BSc ^54^, Dr Sarah Wyllie ^70^ , and Jamie Young BSc ^3^.

**Software and analysis tools**

Amy Gaskin MSc^33^, Dr Will Rowe PhD ^15^, and Dr Igor Siveroni PhD^96^.

**Visualisation:**

Dr Robert Johnson PhD ^96^.

**1** Wellcome Sanger Institute, **2** Public Health England, **3** University of Cambridge, **4** Health Data Research UK, Cambridge, **5** Public Health Agency, Northern Ireland ,**6** Queen's University Belfast **7** Public Health England Colindale, **8** Department of Medicine, University of Cambridge, **9** University of Oxford, **10** Departments of Infectious Diseases and Microbiology, Cambridge University Hospitals NHS Foundation Trust; Cambridge, UK, **11** Division of Virology, Department of Pathology, University of Cambridge, **12** The Francis Crick Institute, **13** Cambridge Institute for Therapeutic Immunology and Infectious Disease, Department of Medicine, **14** Public Health England, Clinical Microbiology and Public Health Laboratory, Cambridge, UK, **15** Institute of Microbiology and Infection, University of Birmingham, **16** University of Birmingham, **17** Queen Elizabeth Hospital, **18** Heartlands Hospital, **19** University of Edinburgh, **20** NHS Lothian, **21** MRC-University of Glasgow Centre for Virus Research, **22** Institute of Biodiversity, Animal Health & Comparative Medicine, University of Glasgow, **23** West of Scotland Specialist Virology Centre, **24** Dept Zoology, University of Oxford, **25** University of Surrey, **26** Wellcome Centre for Human Genetics, Nuffield Department of Medicine, University of Oxford, **27** Big Data Institute, Nuffield Department of Medicine, University of Oxford, **28** Oxford University Hospitals NHS Foundation Trust, **29** Basingstoke Hospital, **30** Centre for Genomic Pathogen Surveillance, University of Oxford, **31** Hampshire Hospitals NHS Foundation Trust, **32** University of Southampton, **33** Public Health Wales NHS Trust, **34** Cardiff University, **35** Betsi Cadwaladr University Health Board, **36** Cardiff and Vale University Health Board, **37** Swansea University, **38** University of Sheffield, **39** Sheffield Teaching Hospitals, **40** Great Ormond Street NHS Foundation Trust, **41** University College London, **42** Oswaldo Cruz Institute, Rio de Janeiro **43** North West London Pathology, **44** Imperial College Healthcare NHS Trust, **45** NIHR Health Protection Research Unit in HCAI and AMR, Imperial College London, **46** University of Liverpool, **47** Manchester University NHS Foundation Trust, **48** Liverpool Clinical Laboratories, **49** University of Exeter, **50** Royal Devon and Exeter NHS Foundation Trust, **51** Quadram Institute Bioscience, University of East Anglia, **52** Norfolk and Norwich University Hospital, **53** University of East Anglia, **54** Deep Seq, School of Life Sciences, Queens Medical Centre, University of Nottingham, **55** Virology, School of Life Sciences, Queens Medical Centre, University of Nottingham, **56** Clinical Microbiology Department, Queens Medical Centre, **57** PathLinks, Northern Lincolnshire & Goole NHS Foundation Trust, **58** Clinical Microbiology, University Hospitals of Leicester NHS Trust, **59** Viapath, **60** Hub for Biotechnology in the Built Environment, Northumbria University, **61** NU-OMICS Northumbria University, **62** Northumbria University, **63** South Tees Hospitals NHS Foundation Trust, **64** North Cumbria Integrated Care NHS Foundation Trust, **65** North Tees and Hartlepool NHS Foundation Trust, **66** Newcastle Hospitals NHS Foundation Trust, **67** County Durham and Darlington NHS Foundation Trust, **68** Centre for Enzyme Innovation, University of Portsmouth, **69** School of Biological Sciences, University of Portsmouth, **70** Portsmouth Hospitals NHS Trust, **71** University of Warwick, **72** University Hospitals Coventry and Warwickshire, **73** Warwick Medical School and Institute of Precision Diagnostics, Pathology, UHCW NHS Trust, **74** Genomics Innovation Unit, Guy's and St. Thomas' NHS Foundation Trust, **75** Centre for Clinical Infection & Diagnostics Research, St. Thomas' Hospital and Kings College London, **76** Department of Infectious Diseases, King's College London, **77** Guy's and St. Thomas’ Hospitals NHS Foundation Trust, **78** Centre for Clinical Infection and Diagnostics Research, Department of Infectious Diseases, Guy's and St Thomas' NHS Foundation Trust, **79** Princess Alexandra Hospital Microbiology Dept. , **80** Cambridge University Hospitals NHS Foundation Trust, **81** East Kent Hospitals University NHS Foundation Trust, **82** University of Kent, **83** Gloucestershire Hospitals NHS Foundation Trust, **84** Department of Microbiology, Kettering General Hospital, **85** National Infection Service, PHE and Leeds Teaching Hospitals Trust, **86** Cambridge Stem Cell Institute, University of Cambridge, **87** Public Health Scotland, 88 Belfast Health & Social Care Trust, **89** Health Services Laboratories, **90** Barking, Havering and Redbridge University Hospitals NHS Trust, **91** Royal Free NHS Trust, **92** Maidstone and Tunbridge Wells NHS Trust, **93** University of Brighton, **94** Kings College London, **95** PHE Heartlands, **96** Imperial College London.
